# Pembrolizumab and Decitabine for Refractory or Relapsed Acute Myeloid Leukemia

**DOI:** 10.1101/2021.06.23.21258377

**Authors:** Meghali Goswami, Gege Gui, Laura W. Dillon, Katherine E. Lindblad, Julie Thompson, Janet Valdez, Dong-Yun Kim, Jack Y. Ghannam, Karolyn A. Oetjen, Christin B. Destefano, Dana M. Smith, Hanna Tekleab, Yuesheng Li, Pradeep K. Dagur, Thomas Hughes, Jennifer L. Marte, Jaydira del Rivero, Joanna Klubo-Gwiezdzinksa, James L. Gulley, Katherine R. Calvo, Catherine Lai, Christopher S. Hourigan

## Abstract

The powerful “graft versus leukemia” effect thought partly responsible for the therapeutic effect of allogeneic hematopoietic cell transplantation in acute myeloid leukemia (AML) provides rationale for investigation of immune-based therapies in this high-risk blood cancer. There is considerable pre-clinical evidence for potential synergy between PD-1 immune checkpoint blockade and the hypomethylating agents already commonly used for this disease. We report here the results of 17-H-0026 (PD-AML, NCT02996474), an investigator sponsored, single-institution, single-arm open-label ten-subject pilot study to test the feasibility of the first-in-human combination of pembrolizumab and decitabine in adult patients with refractory or relapsed AML (R-AML). In this cohort of previously treated patients, this novel combination of anti-PD-1 and hypomethylating therapy was feasible and associated with a best response of stable disease or better in 6 of 10 patients. Considerable immunological changes were identified using TCRβ sequencing as well as single-cell immunophenotypic and RNA expression analyses on sorted CD3+ T cells in patients who developed immune-related adverse events (irAEs) during treatment. Clonal T cell expansions occurred at irAE onset; single-cell sequencing demonstrated that these expanded clones were predominately CD8+ effector memory T cells with high cell surface PD-1 expression and transcriptional profiles indicative of activation and cytotoxicity. In contrast, no such distinctive immune changes were detectable in those experiencing a measurable anti-leukemic response during treatment. Addition of pembrolizumab to ten-day decitabine therapy was clinically feasible in patients with R-AML, with immunological changes from PD-1 blockade observed in patients experiencing irAEs.

**One Sentence Summary:** AML patients receiving a novel combination of a PD-1 immune checkpoint inhibitor with a hypomethylating agent demonstrated clear evidence of induced immunological responses in those developing autoimmune toxicity during treatment but not in those demonstrating an anti-leukemic response.

## Introduction

Acute myeloid leukemia (AML) is an oligoclonal hematological malignancy characterized by myeloid leukemic blasts in the bone marrow (BM) and peripheral blood (PB). The majority of patients diagnosed with AML will develop relapsed or refractory disease (R-AML). Response rates to intensive salvage therapy for AML patients who relapse range from 30-50%, while response rates for those with refractory disease are even lower, typically under 10% (*1, 2*). There is no standard of care for adult R-AML other than referral to an appropriate clinical trial. The graft-versus-leukemia effect of allogeneic hematopoietic cell transplantation for AML provides precedent for the investigation of other immunotherapeutic approaches to treat R-AML. The anti-leukemic benefit of such transplants however is balanced by the risk of severe immunologic toxicity against recipient host tissues (*3, 4*).

Immune checkpoint blockade (ICB) of the PD-1 signaling axis has proven to be one of the most important advances in cancer treatment (*5*). The PD-1 immunoreceptor has two known ligands (PD-L1 and PD-L2); binding of either ligand to PD-1 attenuates T cell receptor (TCR) signaling. Persistent antigenic stimulation can lead to sustained PD-1 expression and T cell dysfunction or exhaustion (*6, 7*). T cell exhaustion is prevalent in many cancers, and therapeutic monoclonal antibodies against PD-1 can abrogate T cell exhaustion and restore functionality (*8-11*).

Seminal murine studies demonstrated a role for PD-1/PD-L1 blockade in the reduction of AML tumor burden (*12, 13*). We and others have shown that CD3+ T cells expressing clinically actionable inhibitory immunoreceptors, including PD-1, are present in the BM and PB of R-AML, even in those with very high burden of disease (*14-19*). Furthermore, there is biological rationale for the combinatorial use of anti-PD-1 agents and hypomethylating agents (HMAs) for R-AML with potential mechanistic synergy between these classes of drugs (*20*). HMAs alone may cause up-regulation of PD-L1/2, leading to resistance and immune escape (*21, 22*). HMAs may also upregulate cancer testis antigens and reactivate previously silenced endogenous retroviruses in leukemic blasts, potentially leading to induction of tumor-specific CD8+ T cells (*23-26*). Importantly though, PD-1 is a critical mediator of peripheral tolerance, as T cells with a degree of autoreactivity upregulate PD-1 to inhibit pathogenic behavior (*27, 28*). Indeed, the association between immune-related adverse events (irAEs) and ICB suggests these agents are subverting normal peripheral tolerance mechanisms, leading to autoimmune toxicities.

Pembrolizumab is a potent and highly selective humanized monoclonal antibody of the IgG4/kappa isotype designed to directly block the interaction between PD-1 and its ligands and reinvigorate existing immune responses in the presence of antigen receptor stimulation. Decitabine is a nucleoside metabolic inhibitor that directly incorporates into DNA and inhibits DNA methyltransferase, causing hypomethylation of DNA and cellular differentiation or apoptosis. Decitabine is commonly used for standard of care treatment of AML (*29, 30*).

Here, we report outcomes of the first clinical trial (NCT02996474) to investigate the novel combination of pembrolizumab and decitabine in R-AML. In addition, to explore potential induced immunological changes during treatment, we used TCRβ DNA-sequencing (TCRβ-seq) on serial samples to construct a picture of clonotypic T cell dynamics during treatment. We then utilized single-cell RNA sequencing (scRNA-seq) with full length VDJ TCR and cell surface antibody-oligonucleotide staining to understand immunogenomic and phenotypic changes associated with the important clinical outcomes of toxicity and efficacy observed in the trial.

## Results

### Safety and efficacy of pembrolizumab with decitabine in R-AML

Ten high-risk previously treated adult R-AML patients (median age 62, range 30-81) were enrolled in the clinical trial 17-H-0026 (PD-AML, 17-H-0026, NCT02996474) between March and November 2017 at the NIH Clinical Center. The treatment schema and research sampling intervals are shown in Fig. 1A. At enrollment seven patients had refractory disease (including two with therapy-related myeloid neoplasms) and three had early relapse within 6 months from completion of last therapy (Fig. 1B). The primary study aim was to determine feasibility of this novel treatment combination; evaluation of safety and efficacy were additional clinical objectives.

**Fig. 1.**
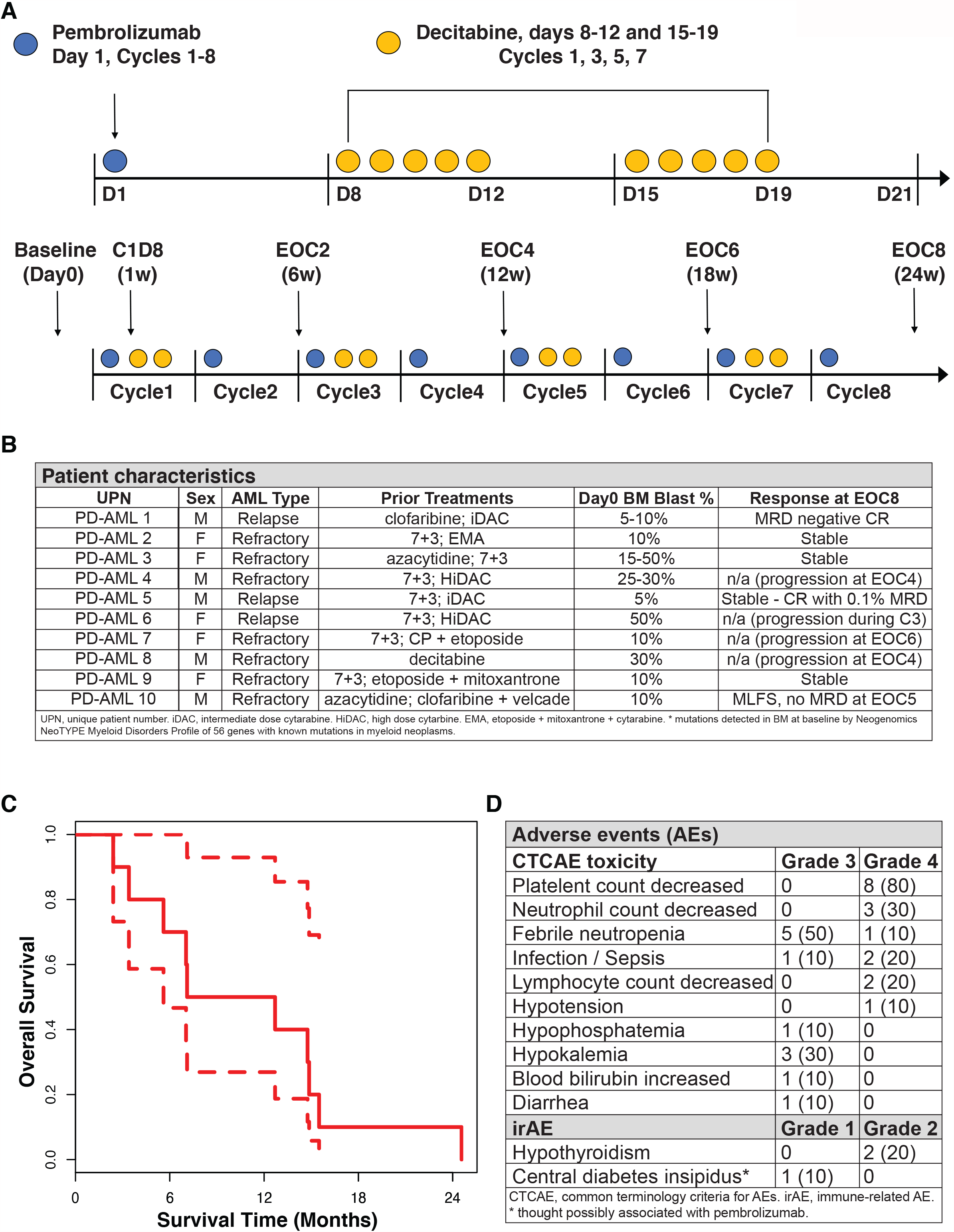
PD-AML clinical trial schema, sampling intervals, and survival curve. (**A**) Pembrolizumab was administered on day 1 of every cycle for 8 cycles, and decitabine was given on days 8-12 and 15-19 of every other cycle. Bone marrow aspirate and peripheral blood was collected before the start of treatment (Day 0), on cycle 1 day 8 (C1D8) after one dose of pembrolizumab, and at the end of every other cycle (EOC2, EOC4, EOC6, EOC8); clinical responses were also assessed at these timepoints. (**B**) Summary of patients treated on NCT02996474 and responses. These patients have been reported previously (*41, 71*). (**C**) Kaplan-Meier survival curve with 95% confidence interval. (**D**) Summary of Grade 3 and 4 adverse events and irAEs that occurred during treatment.

The median number of pembrolizumab doses administered was 6 (mean: 5.8, range 3-8) of a possible 8. The median number of cycles of 10-day decitabine was 3.75 (mean: 3.1, range 1.5-4). The median overall survival from the first day of study treatment was 10 months (mean: 10.8 months, range 2.4 - 24.6 months) (Fig. 1C). No grade 5 adverse events occurred; most grade 4 adverse events were hematological. Non-hematological grade 4 events (sepsis) were seen in two subjects. The toxicity profile of this novel combination therapy was largely consistent with that expected from decitabine alone with the exception of three patients who suffered irAEs likely as a consequence of pembrolizumab (Fig. 1D) This included 2 patients developing hypothyroidism (after 2 and 4 cycles respectively) and a third diagnosed with central diabetes insipidus (after 4 cycles).

In summary, 4 of 10 patients had leukemic progression prior to cycle 8 and were taken off study per protocol rules (Fig. 2). An additional patient was taken off study due to grade 4 infection in cycle 5 while in a morphological leukemic free state (MLFS). For those completing the full planned 24-weeks of the study, 3 had stable disease and 2 were in a cytomorphological complete remission (CR) including one in measurable residual disease (MRD) negative CR (Fig. 2).

**Fig. 2.**
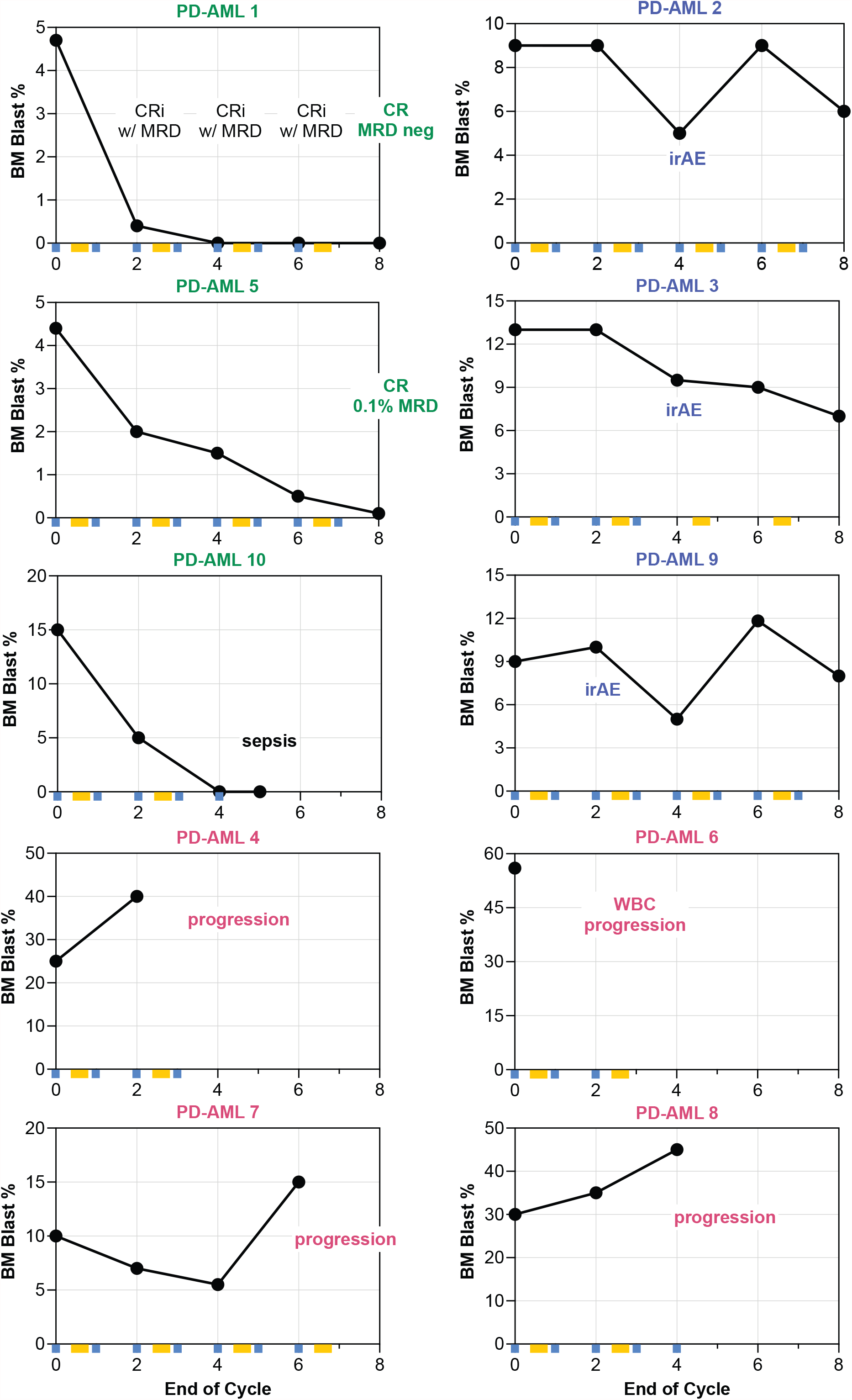
Changes in bone marrow leukemic blast burden in patients treated on NCT02996474. Blast percentages at baseline and after every other cycle of treatment are shown, along with notable clinical events and MRD status, if applicable. Blue squares and along x-axis indicate administered doses of pembrolizumab and yellow rectangles indicate given 10-day dosing of decitabine. Patients with decrease in BM leukemic blast burden (“responsive disease”) are highlighted with green; patients who developed irAEs are indicated in blue, and patients with progressive disease who were taken off study before completion of 8 cycles of treatment are indicated in red.

Specifically, a patient enrolled at second relapse achieved a CR3 lasting 337 days (compared with prior CR2 of 185 days) and was MRD negative by flow cytometry at the end of eight cycles. He survived over 15 months from the start of the protocol treatment despite having significant competing comorbidities. A patient in early first relapse decreased blasts from 4.4% at enrollment to 0.1% at the end of 8 cycles (classified as stable disease, as remained in CR with MRD) and survived over 12 and a half months from the start of protocol treatment.

### Characteristic pattern of clonal T cell expansion in patients developing irAEs

Without a randomized controlled trial, it is challenging to confidently determine if addition of anti-PD1 immunotherapy to clinical standard of care hypomethylating therapy (decitabine) is associated with increased anti-leukemic efficacy. Clinical irAEs however can be confidently attributed to the addition of this immunotherapy, allowing laboratory characterization and quantification of potentiated immune responses.

We first examined abundance and diversity of BM-infiltrating and circulating T cells across all patients and in healthy donors (HD) of similar ages. The proportion of CD3+ T cells in the BM of AML patients prior to treatment did not differ from that of HD, in concordance with prior studies (Fig. S1A) (*14, 15*). We noted an upward trend of CD3+ infiltrate in BM during treatment particularly in responders (Fig. S1B). TCRβ sample clonality, previously reportedly as associated with response to ICB (*31, 32*), as assessed by TCRβ-seq, was also within the ranges observed in HD; there was no association of baseline repertoire clonality or changes in this measure in BM or PB with clinical response during treatment (Fig. S1C-F).

As the etiology and time of clinical diagnosis of irAEs was known, we performed pairwise identification using TCRβ-seq of T cell clones that significantly changed in frequency during treatment compared to baseline in patients developing irAEs. These 3 patients had significant clonal expansions (3-9 clones in BM and 4-15 in PB at the end of 2 cycles of treatment (EOC2), 12-40 in BM and 7-33 in PB at EOC4) (Fig. 3A-B). We asked whether significant expansion of T cell clonotypes from undetectable or near undetectable (<0.1% productive frequency) at baseline were temporally associated with the development of irAEs. Strikingly, we identified several clonally expanded T cells whose expansion coincided with a rise in thyroid stimulating hormone (TSH) and diagnosis of hypothyroidism in patients PD-AML 2 (Fig 3C) and PD-AML 9 (Fig 3D). PD-AML 2 exhibited a period of thyrotoxicosis preceding overt hypothyroidism, previously described with anti-PD-1-induced thyroiditis (*33, 34*). PD-AML 3, who developed central diabetes insipidus after 4 cycles, had pronounced CD8+ and CD4+ clonal T cell expansions that coincided with the irAE and were undetectable prior to Pembrolizumab (Fig 3E). The development of irAEs were preceded by significant albeit temporary expansion of at least one T cell clone in all patients, and patients developing hypothyroidism continued to experience marked clonal T cell expansions one full cycle after irAE onset (Fig. S2). Significantly expanded clones at irAE onset demonstrated concordant magnitudes of expansion in both BM and PB (Fig. S3). These temporal associations between clonal T cell expansion after pembrolizumab and development of irAEs suggest a potential role for T cell mediated immune toxicity.

**Fig. 3.**
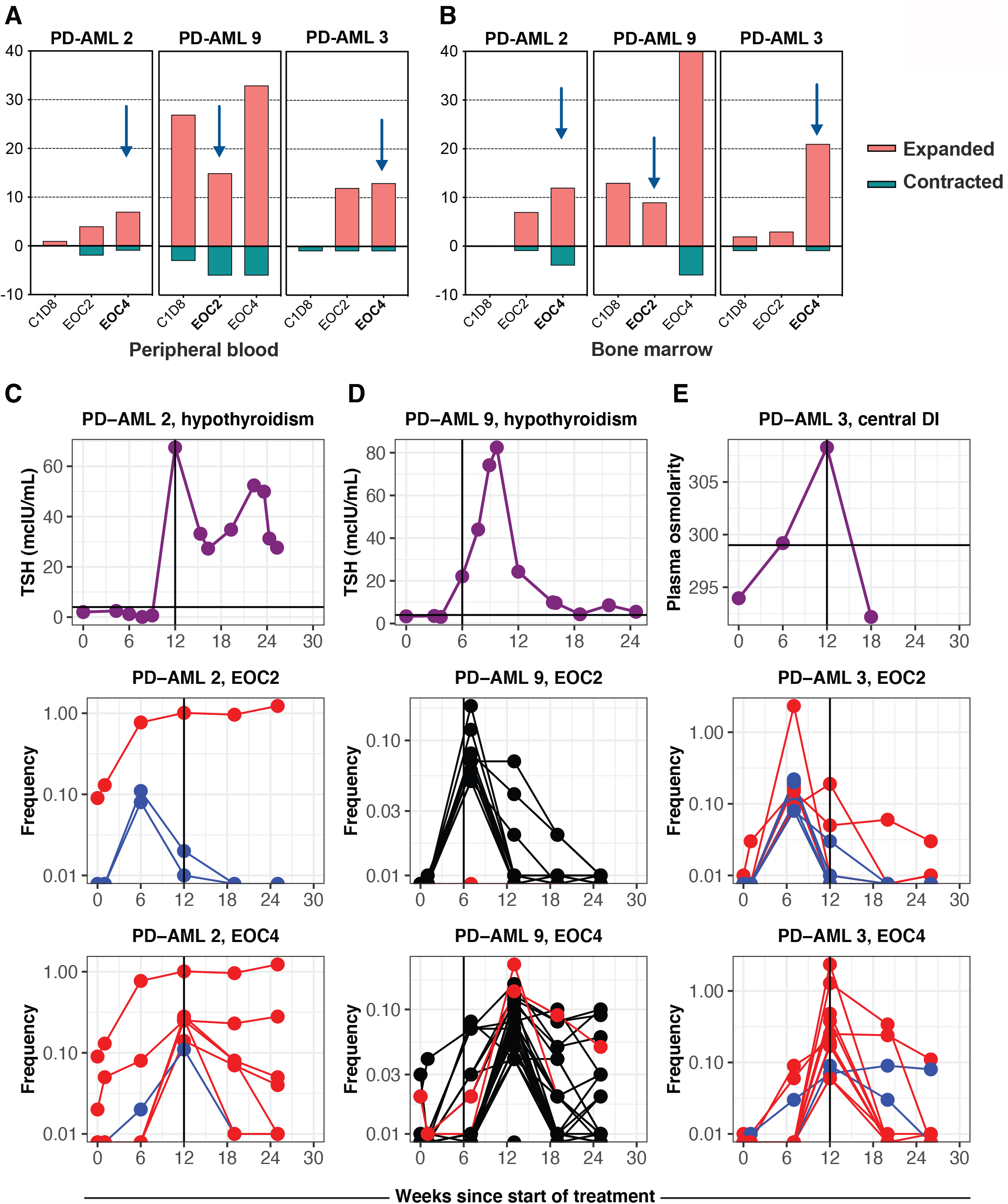
Development of irAEs is associated with clonal T cell expansion. The number of significantly expanded or contracted clones at timepoints sampled during treatment compared to baseline were calculated in (**A**) peripheral blood (PB) and (**B**) bone marrow (BM) for patients developing irAEs. Time of irAE indicated by black arrows. TSH levels and significantly expanded T cells at EOC2 and EOC4 in PB that were <0.1% or undetectable at baseline in (**C**) PD-AML 2, who developed hypothyroidism after 4 cycles of treatment (EOC4, 12 weeks) and (**D**) PD-AML 9, who developed hypothyroidism after 2 cycles of treatment (EOC2, 6 weeks).(**E**) Plasma osmolarity and significantly expanded T cell clones at EOC2 and EOC4 in PB that were <0.1% or undetectable at baseline in PD-AML 3, who developed central diabetes insipidus after 4 cycles of treatment (EOC4, 12 weeks). Upper limit of normal of TSH or plasma osmolarity indicated by black line across y-axes. Each blue, red, or black line represents a unique clonotype for the indicated patient; red indicates CD8+ T cell clones, blue indicates CD8-(CD4+) T cell clones. Black indicates clones where CD4/CD8 could not be determined.

B cells are also capable of mediating autoimmunity, including Hashimoto’s disease (hypothyroidism) (*35*). Previously, we identified a loss of B cells in R-AML patients after chemotherapy (*14*), which was confirmed in this cohort of R-AML patients, including those who developed irAEs (Table S1). At irAE onset, the 2 patients developing hypothyroidism had normal to undetectable autoantibodies against thyroid peroxidase (TPO) and thyroglobulin (TG) (Table S2). In combination, these data suggest a limited role of the humoral immune response in thyroid irAEs in these patients.

### Expanded T cell clones at irAE diagnosis express activation signatures at the transcript and protein level

To understand the distinguishing immunogenomic features of individual T cells clonally expanding during the development of irAEs we next performed 5’ scRNA-seq with full length VDJ TCR sequencing and cell surface antibody-oligonucleotide profiling on FACS-enriched T cells from BM (10x Genomics) from two patients experiencing irAEs (Fig. S4, Table S3).

All CD8+ T cell clonotypes previously identified through bulk TCRβ sequencing of PD-AML 2 at the time of diagnosis of hypothyroidism were also detected by scRNA-seq, along with paired TCRα for most clonotypes of interest. Supervised analysis of irAE-associated CD8+ T cell clonotypes compared with the ten most abundant CD8+ clones identified by TCRβ-seq (all clonotypes had ≥ 5 cells with given CDR3) reveals a striking differentially expressed genes (DEG) profile (Fig. 4A). The majority of the irAE clonotypes expressed granulysin (*Gnly*), indicative of active cytolytic activity, high *Pdcd1* (encoding PD-1 protein), as well as genes related to T cell activation and costimulation (*Sirpg, Psmb8*), survival (*Coro1a, Pycard*), actin remodeling (*Pfn1, Cotla*), and metabolic reprogramming (*Ucp2, Gapdh*). Differentially expressed cell surface proteins (DEP) for the majority of these clones were consistent with an effector phenotype based on CCR7, CD45RA, and CD45RO (Fig. 4B). All expressed very high HLA-DR; the same 4 clones expressing *Pdcd1* transcript co-expressed cell-surface PD-1, TIM-3, and CD27, the latter suggesting recent differentiation from a more memory phenotype (Fig. 4B) (*18*). The 4 clones sharing a common DEG and DEP signature exhibited a similar magnitude expansion in the longitudinal bulk TCRβ-seq data (Fig. 4C). One differentially abundant clone in PD-AML 2 that was detected at baseline highly expressed *Klrc3*, suggesting an invariant NKT-like cell state for this clonotype (Fig. S5A) (*36*).

**Fig. 4.**
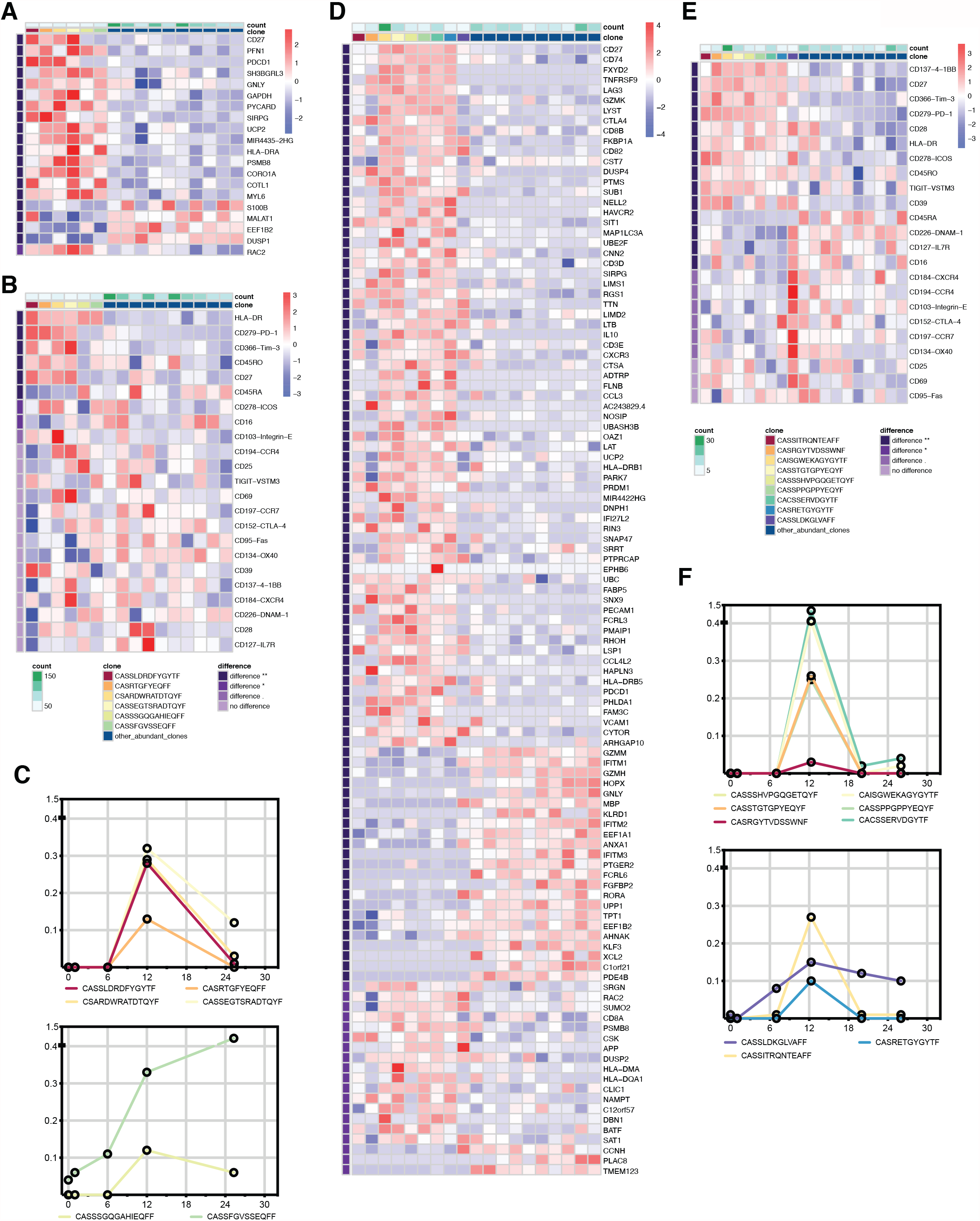
Unique expression signatures in clonally expanded T cells at time of irAE. Cells harboring the unique TCRβ CDR3 from differentially abundant clones identified by bulk TCRβ-seq were grouped and differentially expressed genes (DEG) and cell surface proteins (DEP) were identified by supervised analyses compared to top 10 abundant T cell clones in the sample identified by bulk TCRβ-seq. (**A**) DEG and (**B**) DEP of expanded CD8+ T cell clones of interest in PD-AML 2 at hypothyroidism diagnosis versus other abundant CD8+ T cell clonotypes. (**C**) Frequencies from TCRβ-seq data of clonotypes co-expressing cell surface PD-1, TIM-3, and CD27 (4 clones, top panel) and those not expressing this signature (2 clones, bottom panel). (**D**) DEG and (**E**) DEP of expanded CD8+ T cell clones of interest in PD-AML 3 at time of diagnosis of central diabetes insipidus versus other abundant CD8+ T cell clonotypes. (**F**) Frequencies from TCRβ-seq data of clonotypes co-expressing cell surface PD-1, TIM-3, 4-1BB, HLA-DR, and CD27 (6 clones, top panel) and those not expressing this signature (3 clones, bottom panel). Heatmaps were created using the mean expression of all cells within each clone; differentially expressed markers were ordered by adjusted p-values and fold changes. Adjusted p-values are based on Wilcoxon rank sum test with Bonferroni corrections using all features in the dataset. “difference **” denotes adjusted p-value <0.01; “difference *” denotes adjusted p-value <0.05; “difference.” denotes different but not significantly so. CD8+ T cell clones of interest, expression scale, and significance level are indicated by each heat map. Number of cells in each clone was counted and annotated in green above each heat map.

Using scRNA-seq in the sample acquired at time of central diabetes insipidus diagnosis for PD-AML 3, we detected the majority of expanded clonotypes including both CD8+ and CD4+ T cells. The CD8+ irAE expanded clonotypes were compared to other abundant T cells yielding an even more pronounced DEG profile enriched in T cell activation markers. Six clonotypes highly expressed *CD27, LTB, Lag3, Ctla-4*, and *Havcr2* as well as signature genes identified in PD-AML 2 (*Sirpg, Ucp2*) (Fig. 4D). These same 6 clones highly expressed *Gzmk*, indicating cytolytic capacity. Expanded clones in PD-AML 3 similarly expressed cell-surface PD-1, TIM-3, HLA-DR, and CD27; we noted high 4-1BB expression as well (Fig. 4E). Again, the clones sharing common DEG and DEP signatures largely exhibited similar magnitudes of expansion in the TCRβ-seq data (Fig. 4F). CD4+ irAE expanded clones also were also largely of effector phenotype expressing PD-1, indicative of an activated state (Fig. S5B).

Bulk TCRβ-seq allowed evaluation of a large number of T cells from each timepoint (median: 24,181, range: 13,634-31,247) and hence robust identification of novel and expanded T cell clonotypes, which could then be further characterized using scRNA-seq. In contrast however, the use of just a limited number of T cells using scRNA-seq (Table S3) for discovery of expanded T cell clonotypes would likely lead to false positive results. To demonstrate this, we evaluated T cell clones that were detectable by scRNA-seq at EOC4 but not at earlier timepoints. For both patients, scRNA-seq identified the majority of clones of interest found with bulk TCRβ-seq with the core PD-1, TIM-3, CD27, and HLA-DR cell surface signature evident (Fig. S5C-D). In addition, however, this scRNA-seq based discovery approach falsely identified clones as emerging at EOC4. Bulk TCRβ-seq showed that these clones were most often detectable at baseline or otherwise not significantly expanded, underscoring the limitations of using only scRNA-seq for emerging T cell clone identification (Fig. S5C-D). Downsampling our scRNA-seq datasets to represent experiments performed without pre-enrichment of CD3+ cells by flow cytometry exacerbated this issue as the percentage of overlapping clones between two equal-sized samplings of the same patient sample increased as the number of cells sampled increased (Fig. S6A), indicating higher rates of false-positive clone identification with lower cell numbers. Unique clones from the same patient across timepoints also show little overlap no matter how many cells are sampled (Fig. S6B). These data emphasize the importance of acquired T cell number in scRNA-seq experiments as well as the need for statistical definitions of T cell expansions to limit sampling bias and erroneous identification of novel and emerging T cell clonotypes.

Taken together, the use of bulk TCRβ-seq to accurately profile clonal dynamics followed by T cell enriched scRNA-seq enabled us to characterize at the gene and protein level these significantly expanded T cell clones at time of irAE, revealing their emergence from a memory into effector phenotype with highly activated and cytotoxic immunogenomic signatures.

### Absence of clonal T cell expansions in patients with anti-leukemic responses

Having identified characteristic signatures associated with induced T cell immune responses in patients developing irAEs during treatment with pembrolizumab and decitabine, we next repeated this analysis for patients experiencing an antileukemic response (“responders”). Surprisingly, and in contrast to the above results for patients with irAEs, bulk TCRβ-seq showed that the two patients who completed 24 weeks of therapy and achieved a CR (PD-AML 1 and PD-AML 5) had no significant clonal T cell expansions by EOC4 in either BM or PB (Fig. 5A-B). We found no characteristic clonal expansion associated with decreasing leukemic disease burden during treatment in either patient (Fig. 5C-D). PD-AML 1 developed an infection at EOC6, and the CD8+ expansions seen at this timepoint likely reflect immune responses to this event (Fig. S7A). For PD-AML 5 at EOC6, 2 differentially expanded CD8+ T cell clones were detected that increased in frequency as early as C1D8, although not to statistically significant levels (Fig. S7B). A third responder, PD-AML 10, achieved a MLFS with very hypocellular marrow that precluded reliable estimates of T cell expansions (Fig. S7C). Patients with progressive disease during treatment were either taken off study early or had few clonal T cell expansions (Fig. S8). As done for longitudinal timepoints in two patients developing irAEs, we performed scRNA-seq on baseline, EOC2, and EOC4 timepoints for two responders PD-AML 1 and PD-AML 5. Despite the limitations of novel T cell clone identification through scRNA-seq discussed earlier, given the absence of significant clonal T cell expansions in these responders, we performed supervised analyses with T cell clones detected at both EOC2 and EOC4 that were not found in earlier timepoints. CD8+ T cells in PD-AML 1 at EOC2 and EOC4 that were undetected in prior timepoints by scRNA-seq largely expressed markers indicating activated effector phenotypes, though no shared pattern of PD-1, TIM-3, CD27, and HLA-DR was evident (Fig. S9A). For PD-AML 5 at both EOC2 and EOC4, the number of new T cell clonotypes were so few, identifying meaningful differences compared to previously detected T cell clonotypes was limited (Fig. S9B).

**Fig. 5.**
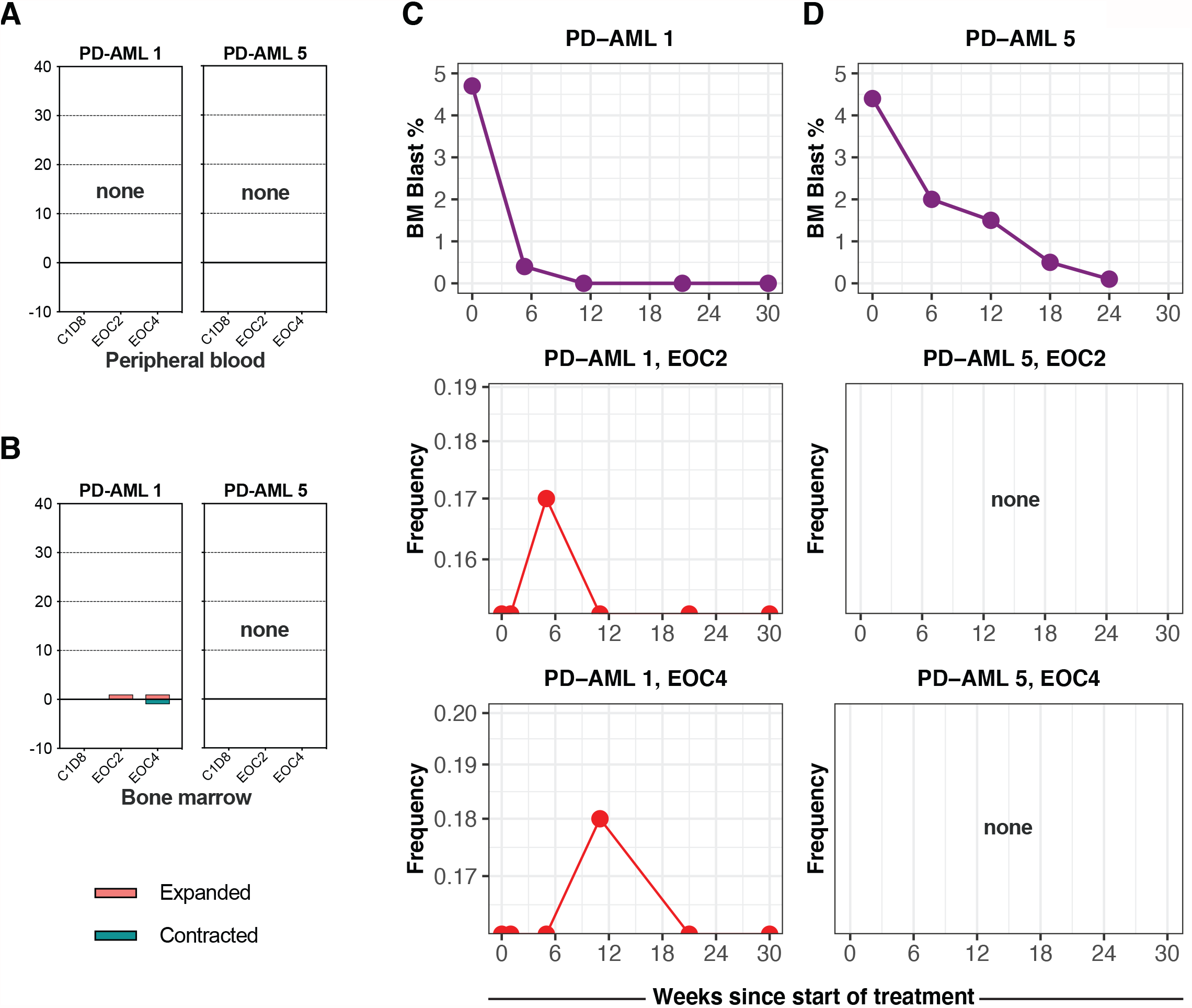
Anti-leukemic response to combination immunotherapy was not associated with clonal T cell expansion. Number of significantly expanded or contracted clones at timepoints sampled during treatment compared to baseline were calculated in (**A**) PB and (**B**) BM for 2 patients achieving CR by 8 cycles of treatment. Decreasing blast percentage in the BM and few to no significantly expanded clones at EOC2 or EOC4 in BM that were low in frequency (<0.1%) or undetectable at baseline in (**B**) PD-AML 1 and (**C**) PD-AML 5. Red lines represent unique CD8+ clonotypes.

### No changes in T cell immunophenotypes and expression profiles in responders during treatment

An alternative hypothesis would be that pembrolizumab would not lead to detectable clonal expansions at the tumor site, but instead would change the activation status of already resident T cell clones. To investigate this possibility, we shifted from clone-driven inquiries to immunophenotype-based analyses. The inclusion of oligonucleotide-labeled antibodies in scRNA-seq allowed for immunophenotyping in a manner analogous to flow cytometry. Using T cell subset-defining markers, we identified CD4+ and CD8+ naïve, central memory (CM), CD4+ CD127lo CD25hi regulatory T cells (Treg), effector memory (EM), and EM re-expressing CD45RA (terminal effectors, or TE) T cell populations (Fig. 6A-B, Fig. S10). Frequencies of CD4 and CD8 T cells derived from scRNA-seq matched well with frequencies calculated from clinical flow cytometry of fresh BM aspirate (Fig. S11A). We confirmed expression of canonical genes that define these given subsets, demonstrating concordance between transcriptional and cell surface expression profiles (Fig. 6C).

**Fig. 6.**
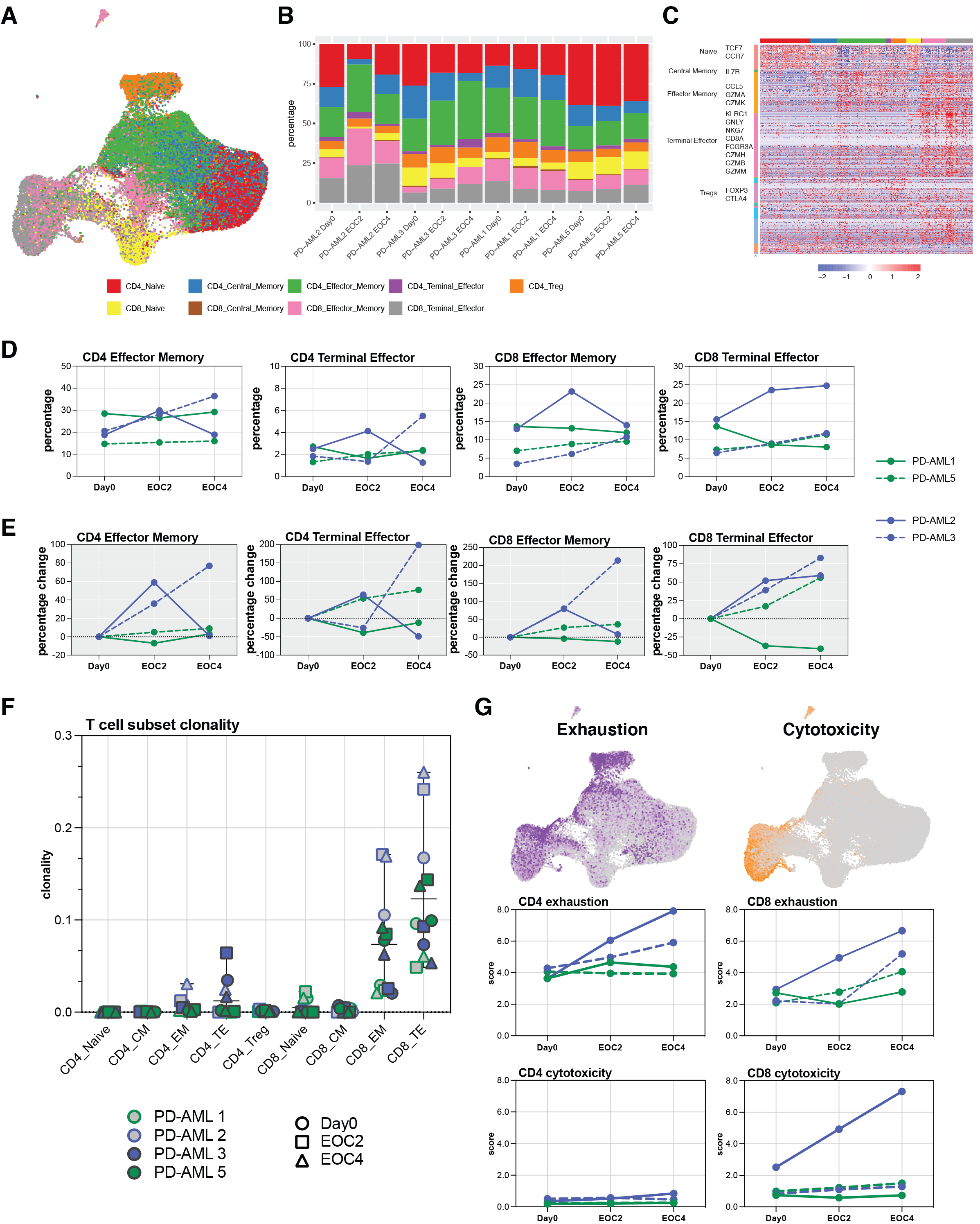
Stable T cell immunophenotypes in patients with anti-leukemic responses. **(A)** Integrated UMAP visualization of T cell immunophenotyping based on cell surface expression CD4, CD8, CCR7, CD45RA, CD127 and CD25. (**B**) Quantification and proportion of CD4 and CD8 T cell subset frequencies, as a percentage of CD3+. (**C**) Differential gene expression analysis among different cell types with scaled log normalization. For panels A-C, T cell subsets indicated in legend. (**D**) Frequencies of CD4+ and CD8+ effector memory (EM) and terminal effector (TE) T cells at Day0, EOC2, and EOC4. (**E**) Percent change in frequencies of CD4+ and CD8+ EM and TE cells at EOC2 and EOC4 compared to Day0. (**F**) T cell subset clonality in T cell subsets identified by immunophenotyping. Patients and timepoints indicated in key. (**G**) Cells expressing exhaustion (purple) and cytotoxicity (orange) predefined gene sets overlaid on integrated UMAP with exhaustion and cytotoxicity scores for CD4+ and CD8+ T cells. Scores were calculated for each cell based on mean expression of genes in the gene set and subtracting the background. For panels D-F, individual patients indicated in legend.

Large changes in frequencies of effector T cell subpopulations were seen in patients developing irAEs, with these patients experiencing at EOC2 36-59% increases in CD4 EM and an 80% increase in CD8 EM (Fig. 6D-E). Both patients had 39-52% increases in CD8 TE at EOC4 as well. Responders maintained stable frequencies of EM populations, though PD-AML 5 experienced over 50% increase in both CD4+ and CD8+ TE populations by EOC4 (Fig. 6D-E). Patients developing irAEs had decreases in frequencies of naïve CD4+ and CD8+T cells during treatment; all patients had slight decreases in Treg frequencies by EOC4 (Fig. S11B-C). We derived overall sample clonality for T cells sequenced with scRNA-seq, which had excellent concordance with bulk clonality values obtained with bulk TCRβ-seq (Fig. S11D-E). Our workflow enabled us to compute sample clonality for these specific T cell subsets as well. The increases in CD8 EM frequencies in patients developing irAEs were accompanied by increases in clonality (Fig. 6F), likely a reflection of the clonal expansions we identified earlier. CD8 TE clonality was also increased in PD-AML 5 at EOC4, suggesting increase in frequency of a more limited number of pre-existing CD8 T cell clones. Overall, however, responders maintained diverse CD4 and CD8 subpopulation TCRβ repertoires through EOC4.

The maintenance of high frequency clones that persist from baseline may be associated with response to ICB (*37*). The top 10 most abundant clones identified by TCRβ-seq for all patients at baseline were consistent throughout subsequent sampled timepoints and all fell within CD8 EM and TE clusters (Fig. S12A-B). We next investigated whether the phenotype of these consistently abundant clones changed with treatment. While these abundant clones had differing cell surface profiles from one another, their phenotypes at EOC2 and EOC4 remained unchanged from baseline (Fig. S12C). We also searched for clonotypes in the responders that may have experienced shifts from memory to more effector and/or terminal phenotypes. While we observed heterogeneity in phenotypic composition within clonotypes, the profiles of the clonotypes at baseline were maintained throughout treatment. We found no clonotype with uniform shifts between T cell phenotypes (Fig. S12D).

Finally, we derived naïve differentiation state, exhaustion, and cytotoxicity scores by computing average scores of gene modules associated with these cell states (*38*). Of note, cells expressing genes within a cytotoxic module were almost exclusively CD8+ effector and terminal effector phenotypes, while cells expressing exhaustion modules consisted of all CD4 and CD8 T cell subsets (Fig. 6G, top). At baseline, all 4 patients had similar CD4 exhaustion and cytotoxicity scores; at EOC4, patients who developed irAEs had averages of 0.78-fold and 1.31-fold in CD4 and CD8 exhaustion scores, respectively (Fig. 6G). PD-AML 2 also demonstrated a sharp increase in CD8 cytotoxic state at EOC4; these data are consistent with the clinical development of irAEs at this timepoint in both patients and cellular profiles described earlier. PD-AML 5 demonstrated an increase in CD8 exhausted state at EOC4, and PD-AML 1 maintained low exhaustion and cytotoxicity scores through treatment (Fig. 6G). These data suggest increases in aggregate cell cytotoxicity scores are associated with the development of irAEs rather than anti-leukemic response to treatment in these patients.

### No evidence of HLA loss in responders at relapse

A third approach to test the hypothesis that anti-PD1 immunotherapy was associated with observed anti-leukemic efficacy is the evaluation for potential immune escape at the time of relapse in those previously responding. Despite continuing maintenance therapy after the completion of this 24-week trial in CR, PD-AML 1 and 5 ultimately both relapsed 374 and 221 days after initiation of protocol treatment, respectively. HLA downregulation or loss in AML has been reported at relapse after allogeneic transplant (*39, 40*). We therefore performed both sc-RNA-seq and single cell DNA sequencing (scDNA-seq) profiling at baseline and relapse timepoints with unsorted bone marrow mononuclear cells (BMMCs) from these two patients to evaluate for any evidence of cell-surface HLA loss on the leukemic clone as evidence of immune selective pressure.

Clustering based on cell surface protein expression of scRNA-seq (10x Genomics 3’ v3) identified the full array of expected immune cells within each unsorted bone marrow sample (Fig. S13). We focused on populations expressing leukemia-associated immunophenotypes and quantified relative expression of HLA class I and II as well as PD-L1 expression on these putative leukemic clusters. In both patients at relapse, leukemic clusters had stable HLA class I (A, B, C) and class II (DR, DQ, DP) at relapse compared to baseline (Fig. 7A-B). PD-L1 expression was not increased at relapse for either patient. As use of cell-surface immunophenotype may imperfectly identify leukemic cell populations, we confirmed these findings with an alternative approach. We have previously reported the use of patient-personalized scDNA-seq on baseline samples for these two patients to resolve leukemic clonal landscape (*41*). We therefore also acquired scDNA-seq with oligonucleotide-conjugated antibodies against myeloid markers on these relapse samples (Fig 7C-D). We confirmed that the relapsing AML had the same genomic mutational features and cell-surface immunophenotype as baseline and that the HLA class I and class II of the leukemic clone remained stable at relapse compared with initial diagnosis.

**Fig. 7.**
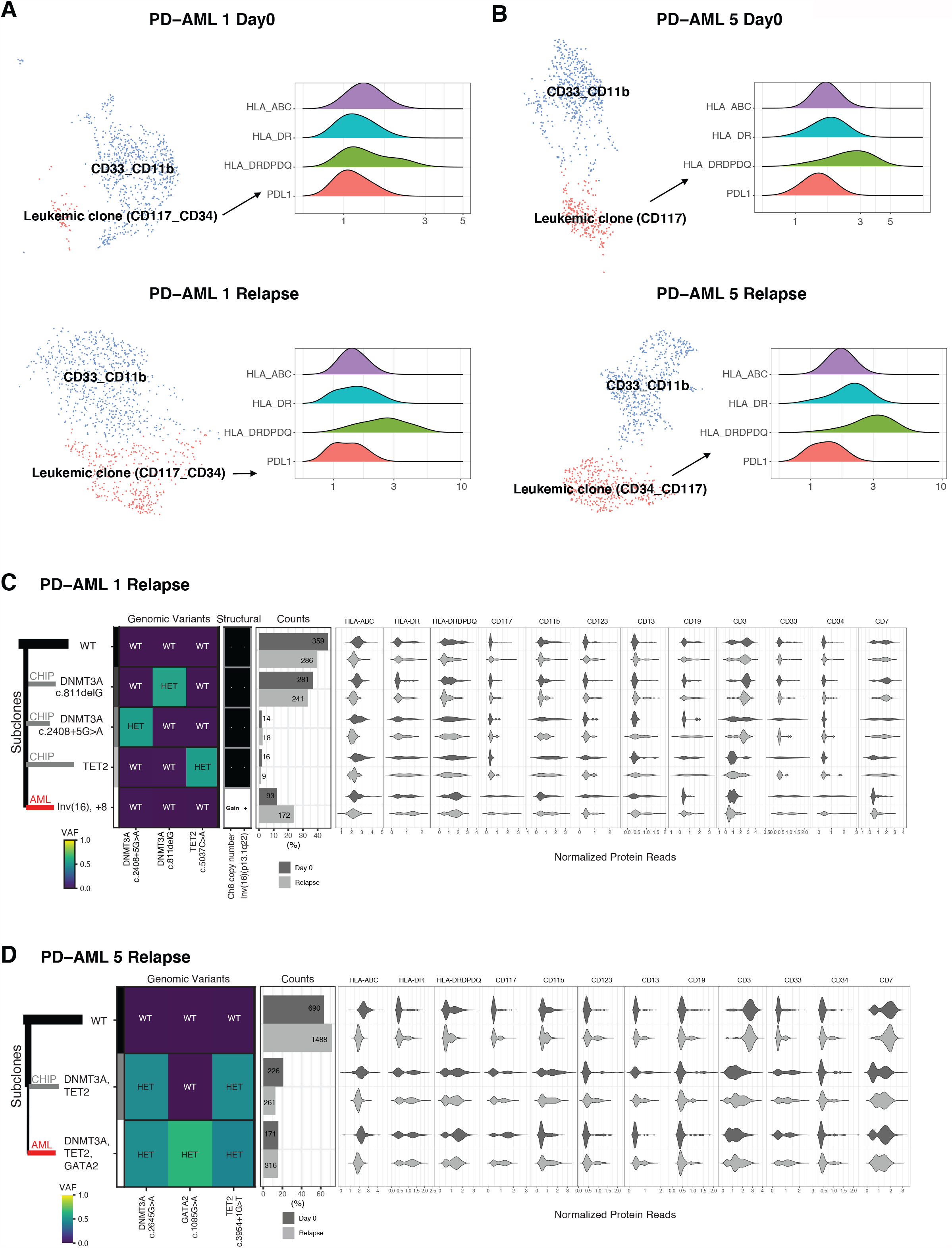
No signal of immune evasion in responders at relapse post-treatment. Through 3’v3 scRNA-seq on unsorted BMMCs, clustering was based on the cell surface protein expression and clusters were annotated by their top 3 or 4 most highly differentially expressed markers. Clusters were classified as putative leukemia using on protein expression of known leukemia-associated and myeloid markers based on the patients’ previous clinical flow cytometry records. Relative expression of HLA molecules and PD-L1 on leukemic blasts at baseline (top) and at relapse (bottom) in (**A**) PD-AML 1 and (**B**) PD-AML 5 visualized using ridge plots. Single-cell DNA and antibody-oligonucleotide sequencing of unsorted BMMCs in (**C**) PD-AML 1 and (**D**) PD-AML 5 shows the genomic and immunophenotypic features of the leukemic (AML, red line) and preceding/distinct CHIP (clonal hematopoiesis, grey lines) clones present at relapse as compared to Day 0 (adapted from *(41)*). Left: Genomic subclones with wild type (WT), heterozygous (HET), present (Gain/+) or absent (-) features. Right: Cell surface protein expression for each subclone.

## Discussion

This trial is the first-in-human study of pembrolizumab and decitabine to treat R-AML. We performed integrated multi-dimensional immunological analysis including longitudinal paired TCRβ measurements in PB and BM (i.e.: the sites of tumor), together with scRNA-seq with full length VDJ TCR sequencing and cell surface antibody-oligonucleotide profiling of isolated T cells to further our understanding of how this immunotherapeutic combination impacts T cell dynamics in R-AML.

While the preclinical rationale for the addition of PD-1 axis ICB to hypomethylating therapy for the treatment myeloid malignancies is strong (*21-24*), the clinical evidence base for improved efficacy is lacking. One other study to date has reported clinical results in R-AML patients treated with a PD-1 inhibitor in conjunction with a hypomethylating agent, with a similar overall response rate (33%) to that observed here (30%) (*42*). No clinically meaningful difference in efficacy was noted in a randomized, open-label, international, multicenter study that enrolled untreated myelodysplastic syndrome (MDS) and AML patients to either azacitidine and the anti-PD-L1 agent durvalumab versus azacitidine alone (*43*). Of note, the two patients completing the 24 weeks of planned therapy in CR entered our study with relapsed, MRD level of disease between 0% and 5% blasts. Currently recruiting are randomized studies focus on the addition of pembrolizumab for MRD in combination with either intensive chemotherapy (NCT04214249) or azacitidine and venetoclax (NCT04284787), which will offer better idea of efficacy of ICB particularly in the MRD disease level setting.

Notable clinical events in the present trial were irAEs in 3 (30%) patients, affecting endocrine organs. Meta-analyses suggest that as many as 20% of patients treated with pembrolizumab will develop a thyroid-related autoimmune condition, as was our observation here (*44*). Central diabetes insipidus is a rare irAE affecting the posterior pituitary; one other case of central diabetes insipidus has been reported in a patient treated with the PD-L1 inhibitor avelumab (*45*). A phase I trial of nivolumab as a maintenance therapy post allo-HSCT in patients with AML or MDS (NCT02985554) recently closed early after only 4 patients were treated because of the severe irAEs that occurred in all 4 patients, including one case of hypothyroidism (*46*). The use of nivolumab and ipilimumab either in combination or sequentially is ongoing in AML patients after allo-HSCT (NCT03600155), though no results have been released to date. The relationship between irAE development and anti-tumor response in AML remains unknown (*47*).

A key finding of this study comes from the in-depth statistical analyses of specific T cell clonal frequencies during the course of treatment. Differential clonotypic abundance analyses revealed distinctive clonal expansions from previously undetectable levels at the onset/diagnoses of irAEs in 3 patients treated with pembrolizumab and decitabine. Large numbers of clonal CD8+ T cell expansions preceding the development of grade 2-3 irAEs and early diversification in peripheral TCRβ repertoire in patients who ultimately developed irAEs during treatment with ipilimumab have been reported (*48, 49*). Our findings differ, as we did not observe broad repertoire shifts in terms of clonality or clonal T cell expansions until a minimum of 6 weeks after pembrolizumab initiation in our R-AML patients developing irAEs. These differences in clonal kinetics may reflect the different mechanisms of actions of anti-CTLA4 and anti-PD-1 drugs (*11*). Infiltration of memory cytotoxic CD4+ and CD8+ T cells has been identified in ICB-induced encephalitis, and recent single cell studies in patients developing colitis irAEs during ICB identify signatures of proliferation and IFNγ responsiveness in colitis-associated T cells (*50, 51*). Our findings of PD-1 and HLA-DR expression on expanded clones of interest are consistent with these reports. We further identify a shared expression module of these two markers as well as TIM-3 and CD27, along with a transcriptional profile indicative of cytotoxicity, cell activation, and co-stimulation in irAE-associated T cells. Based solely on the cell surface co-expression of PD-1 and TIM-3 on these CD8+ T cells, we may have described many of these expanded clones as exhausted or dysfunctional (*52, 53*). However, their transcriptional activity and temporal association with irAEs suggest otherwise. Indeed, a strength of our approach is through enrichment of CD3 T cells for scRNA-seq, which allowed for orthogonal validation and characterization of clones found through bulk TCRβ-seq and integration of temporal dynamics with immunogenomics of significantly expanded T cell clones.

Determining immune correlates of anti-leukemic response in AML patients to ICB is challenging in the absence of randomized clinical trial data (*54, 55*). In the current study for two patients who experienced anti-leukemic benefit during treatment, we intensively scrutinized many CD8+ cellular determinants, including co-expression of inhibitory receptors and activation markers with PD-1, differentiation state of infiltrating T cells with treatment, as well as bystander T cells, all of which have been associated with response to ICB in various solid tumors (*56-59*). However, we found no distinctive changes in these responders and show in fact that instead, changes in these cellular determinants occurred in patients developing irAEs. Moreover, we show here, using the positive control of patients experiencing irAEs clearly attributable to ICB, that assessment of apparently novel or expanded T cell clonotypes by single cell sequencing is subject to sampling bias and false discovery.

The small size of our R-AML cohort is a limitation, as this was a feasibility pilot trial not powered for clinical efficacy assessment. Though some immunomodulatory activities of HMAs on various immune cells have been described, with our trial design, we cannot separate out immunological phenomena caused by pembrolizumab or decitabine alone (*20, 60*). Several studies suggest blocking PD-1 may not reinvigorate exhausted and silenced CD8+ T cells due to extensive and non-reversible epigenetic remodeling (*61, 62*), though we did not investigate the pre-and post-treatment epigenetic landscape in the current study. Future work will investigate the effect of HMAs alone at the single cell level on AML as well as infiltrating and circulating immune cells for better understanding of their immunomodulatory consequences and anti-leukemic activities. We did not find evidence of T cell-mediated anti-leukemic responses in the two responders, but we cannot rule out roles of other immune cells. A recent study suggests that response to anti-PD-1 therapy may be dependent on PD-1 expression and signaling in cells of myeloid origin (*63*). The role of myeloid cells in response to ICB is further supported by a study in advanced stage melanoma patients that identified the starting frequency of CD14+ HLA-DRhi monocytes as a strong predictor to response to treatment with anti-PD-1 agents (*64*). Going forward it will be important to investigate expression of immune checkpoints in non-T cellular compartments in AML both before and during treatment, as their expression on myeloid cells cohabitating with leukemic blasts could influence outcomes during treatment with ICB.

This work is widely relevant to understanding mechanisms behind both irAEs and anti-tumor responses during ICB. Our methods together uncovered significant expansions of activated effector memory CD8+ T clones that occurred in a subset of patients developing autoimmune toxicities during treatment with pembrolizumab and decitabine. The combination of immunogenomic signatures and temporal association with irAEs suggest that these clones were previously peripherally silenced and then stimulated during treatment with ICB. Notably, these signatures of T cell expansion and activation were not evident in two patients with relapsed AML who had anti-leukemic responses to treatment. Our findings contribute to the understanding of immune underpinnings of irAEs induced by ICB and these data serve as a valuable resource for examining the effects of pembrolizumab and decitabine in combination on T cell dynamics in AML at the single cell level.

## Materials and Methods

### Clinical Trial

17-H-0026, (PD-AML, NCT02996474) was an investigator sponsored, single-institution, single-arm open-label ten subject study approved by the NHLBI IRB and conducted in accordance with the Declaration of Helsinki (FDA IND: 131826). Pembrolizumab 200 milligrams was administered intravenously on day 1 of every three-week cycle, with decitabine 20 milligrams per meter squared administered on days 8-12 and 15-19 (i.e.: total of 10 days) of alternative cycles starting with cycle 1. Up to eight cycles (24 weeks) of therapy were given, with the opportunity for an optional continuation phase for those who maintained a best response of at least stable disease after the initial planned eight cycles. Patients were adults with an unequivocal diagnosis of relapsed or refractory acute myeloid leukemia confirmed by an NIH attending pathologist within 30 days of study enrollment. Those with acute promyelocytic leukemia, prior allogeneic hematopoietic stem cell transplant, active autoimmunity or second malignancy, prior PD-1 axis inhibitor therapy or more than two cycles of prior decitabine therapy were excluded from participation. Full eligibility, response and discontinuation criteria are reported in Tables S4 and S5. BM examinations for clinical response assessment were performed prior to treatment and after cycles 2, 4, 6 and 8 (or the progression/off-study time-point). An additional bone marrow examination was performed for research purposes on day 8 of the first cycle (i.e., one week after the first dose of pembrolizumab, immediately before the initiation of decitabine).

### Sample Collection

Bone marrow (BM) aspirate and peripheral blood (PB) was collected from enrolled patients. A cohort of healthy donors (HD, n=13) was also recruited as a control population at the NIH; we previously intensively charactered the BM of this cohort with scRNA-seq, flow cytometry, and immunohistochemistry of BM core biopsies (*15, 65*). This research was approved by the NHLBI IRB, and all patients and HD provided oral and written informed consent under IRB-approved protocols conducted in accordance with the Declaration of Helsinki. BM mononuclear cells (BMMCs) were purified from BM aspirate using density centrifugation, cryopreserved, and stored in liquid nitrogen. CD8+ T cells were enriched from thawed BMMCs with CD8 positive microbeads according to manufacturer’s protocol (Miltenyi Biotec).

### Nucleic Acid Isolation

Genomic DNA (gDNA) was isolated from 2ml of fresh heparinized BM and 2ml of PB using DNA Blood Midi Kits (Qiagen) according to manufacturer’s instructions. AllPrep RNA/DNA Mini Kits (Qiagen) were used to isolate gDNA from CD8+ T cell populations enriched from BMMCs. All gDNA was quantitated using both absorbance (NanoDrop, Thermo Fisher) and fluorometric (Qubit, Thermo Fisher) methods.

### Bulk TCRβ sequencing and T cell receptor data analyses

The CDR3 region of rearranged TCRβ genes was sequenced using the survey depth immunosequencing platform from Adaptive Biotechnologies. Target gDNA input from BM and PB samples was 1-2μg, and the entire gDNA yield from sorted CD8+ T cells was input into the sequencing assay. Amplification and sequencing of the TCRβ CDR3 was performed according to previously described protocols (*66, 67*), and sequenced regions were filtered, mapped, and defined according to the IMGT database (*68*). The ImmunoSEQ toolset from Adaptive Biotechnologies was used to explore and analyze TCRβ sequencing datasets. Significantly differentially abundant T cell clones between two timepoints for a given patient were identified by employing Fisher exact statistical tests fitted with a betabinomial probability density model to account for normal variance in TCRβ repertoires (*69, 70*). T cell clones were considered differentially abundant if their Benjamini-Hochberg (BH)-corrected false discovery rate (FDR) is less than 0.01.

### Cell surface staining with oligo-tagged antibodies and enrichment of CD3+ T cells

To purify CD3+ T cells from cryopreserved BMMCs from AML patients, frozen cells were thawed into warmed 37°C RPMI-1640 supplemented with 50% FBS, washed twice, and counted with trypan blue staining on an automated hemocytometer. Between 500,000 to 1 million cells were resuspended in 56ul of RPMI-1640 with 10% FBS, to which 5ul of TruStain FcX and 2ul of a 1:1 weight to volume solution of dextran sulfate was added. Cells were incubated with the FcX block and dextran for 10 minutes at room temperature (RT), after which 10ul of FITC-conjugated anti-human CD3 (clone HIT3a, Biolegend) and 0.5ug each of the 27 TotalSeq-C oligo-tagged antibodies (Biolegend) listed in Table S6 were added. Cells were incubated with antibodies for 30 minutes at RT in the dark. After incubation, cells were washed twice with RPMI-1640 with 10% FBS and resuspended in 300ul of the same buffer. Five minutes prior to the start of FACS acquisition, 2ul of a 1mg/ml DAPI live dead stain (Thermo Fisher Scientific) was added to the cell suspension, and cells were filtered through 40um cell strainers immediately prior to acquisition. FACS was performed using a BD Aria II equipped with Diva software. After debris and doublet exclusion, live CD3+ T cells were sorted into RPMI-1640 with 10% FBS using a 100um nozzle instead of the standard 20um nozzle, reducing the pressure per cell from 70psi to 20psi during the sorting process. A minimum of 10,000 live T cells were sorted. Immediately after sorting, samples were centrifuged at 250xg for 5 minutes using a swing-arm rotor centrifuge, and volume was adjusted so estimated cell concentration based on cell number sorted was 1000-2000 cells/ul. Cell concentration was verified using trypan blue stain on an automated hemocytometer.

### 5’ Single cell RNA-sequencing (scRNA-seq) library construction and sequencing

The 10x Genomics 5’ Single Cell Immune profiling platform was used for scRNA-seq of enriched T cells. After FACS of oligo-stained CD3+ T cells, the cells were loaded on a Chromium Chip with master mix, gel beads, and partitioning oil according to manufacturer’s instructions. Intended number of cells captured was 6000-8000. Reverse transcription (RT) of mRNA into barcoded first strand cDNA, as well as construction of gene expression (GEX), VDJ, and antibody-derived tag (ADT) sequencing libraries was performed according to the experimental protocol and reagents provided by 10x Genomics, without modifications. GEX, VDJ, and ADT libraries were evaluated using an Agilent Tapestation and quantitated with qPCR (Kapa). The scRNAseq libraries were pooled and sequenced with an Illumina NovaSeq S4 flow cell using an Xp loader. Libraries were sequenced PE28-2-98 with 1% PhiX. Target sequencing depth for the GEX libraries was 20,000 read pairs per cell and 5,000 read pairs per cell for the VDJ and ADT libraries.

### 5’ scRNA-seq Data Processing and Analyses

5’ sequencing libraries were preprocessed using Cell Ranger version 3.1.0 (10x Genomics) to obtain gene expression, feature barcoding counts, and VDJ information. The number of barcode mismatches was set to be 0 to minimize the demultiplexing error. Seurat v3 was used across the study for sample basic quality control, normalization, data integration, visualization, and differential expression. Default methods were used unless otherwise specified for specific tasks. Data from all samples were integrated for visualization using the standard workflow on Seurat 3 by identifying the anchors between individual samples using the top 2000 variable genes within each and reducing dimensions by principal component analyses (PCA) then uniform manifold approximation and projection (UMAP). Immunophenotyping of the cells was analogous to flow cytometry where raw counts of marker proteins including CD3, CD45, CD4, CD8, CD127, CD25, CCR7, and CD45RA were plotted and consensus cutoffs were created by comparing median cutoffs across different timepoints within one sample and adjusting them across all patients.

### 3’ Single cell RNA-sequencing (scRNA-seq) library construction and sequencing

The 10x Genomics 3’v3 Single Cell Immune profiling platform was used for scRNA-seq of BMMCs. Frozen cells were thawed as described earlier, with the addition of a wash step with 5ml of ACK lysis buffer to lyse any residual red blood cells. Up to 1 million cells were resuspended in 50ul of cell labeling buffer (PBS + 1% BSA). Cells were incubated with TruStain FcX (Biolegend) for 10min at 4C, after which 0.5ug each of the 46 TotalSeq-A oligo-tagged antibodies (Biolegend) listed in Table S7 were added. Cells were incubated with antibodies for 30 minutes at RT at 4C. After incubation, cells were washed three times with cell labeling buffer and filtered with 40um Flowmi cell strainers. Final volumes were adjusted so estimated cell concentration was 700-1200 cells/ul using trypan blue stain on an automated hemocytometer. Oligo-tagged cells were loaded on a Chromium Chip B with master mix, gel beads, and partitioning oil, again according to 10x Genomics instructions. cDNA amplification was performed according to manufacturer’s instructions with the addition of 1ul 0.2uM ADT additive primer (Table S8) per sample to amply ADT tags. During cDNA cleanup, supernatant containing the ADT cDNA fraction was kept and used for generation of ADT libraries. Two rounds of 2X ADT cDNA purification were performed with SPRIselect beads, and 100ul PCR reactions for library preparation and amplification were prepared by mixing 2.5ul of purified ADT cDNA fraction with 2.5ul of 10uM SI PCR primer, 2.5ul 10uM Illumina TruSeq small RNA RPI primer, 50ul 2X Kapa Hifi PCR master mix, and 50ul RNAse-free water. All primer sequences for ADT library generation are detailed in Table S9. Reactions were amplified in a thermocycler at 98C for 2 mins, then 14 cycles of 98C for 20sec, 60C for 30sec, and 72C for 20sec, followed by 5min at 72C. The amplified ADT libraries were purified with 1.2X SPRIselect beads. 3’v3 GEX libraries were constructed exactly according to 10x Genomics protocol. 3’v3 GEX and ADT libraries were evaluated as earlier described, and scRNAseq libraries were pooled and sequenced with an Illumina NovaSeq SP flow cell using an Xp loader. Libraries were sequenced PE28-2-91 with 1% PhiX. Target sequencing depth for the GEX libraries was 20,000 read pairs per cell and 5,000 read pairs per cell for the ADT libraries.

### 3’v3 scRNA-seq Data Processing and Analyses

The standard pipeline for Cell Ranger version 3.1.0 (10x Genomics) was used to obtain UMI counts for gene expression and cell surface antibodies. The number of barcode mismatches was set to 0 to minimize demultiplexing error. Seurat v3 was used for sample basic quality control, normalization, visualization, and differential expression. Each sample was analyzed individually, and clustering was performed based on normalized cell surface protein expression. Highly expressed proteins were identified for cell type annotation and leukemia associated clusters were labeled based on markers from previous clinical flow cytometry records.

### Single cell DNA-sequencing (scDNA-seq) library construction, sequencing, and analysis

The MissionBio Tapestri scDNA-seq V2 platform with antibody-oligonucleotide staining was performed on BMMCs per manufacturer’s protocol. Custom primers targeting patient-specific mutations and chromosomal abnormalities, as well as oligonucleotide antibodies against CD45, CD34, CD117, CD11b, CD123, CD13, CD33, CD38, CD90, CD7, CD3, CD19, HLA-A,B,C, HLA-DR, and HLA-DR,DP,DQ were used in this assay. All experimental acquisition, library generation, sequencing, data processing, and analyses were performed exactly as previously described (*41*).

### Statistical Analyses

GraphPad Prism (version 8.4.3) and R 4.0.0 were used for statistical analyses and graphing. Several R packages (ggplot2, Seurat, pheatmap) were used for data manipulation and visualization. This work utilized the computational resources of the NIH HPC Biowulf cluster. (http://hpc.nih.gov).

## Data Availability

Bulk TCRb sequencing data is available on the ImmunoSEQ Analyzer through Adaptive Biotechnologies. scRNA-seq data sets are available through the National Center for Biotechnology Informations Gene Expression Omnibus. Code available upon request.

## Acknowledgements

We appreciate the technical expertise of the NHLBI Flow Cytometry and DNA Sequencing Cores. Thank you to Steven Soldin (NIH Clinical Center) for technical guidance. We thank the NIH HPC Biowulf cluster for their computational resources (http://hpc.nih.gov). This work is supported in part by a research grant from Investigator-Initiated Studies Program of Merck Sharp & Dohme Corp. The opinions expressed in this paper are those of the authors and do not necessarily represent those of Merck Sharp & Dohme Corp.

## Funding

This work was supported by the Intramural Research Program of the National Heart, Lung, and Blood Institute (NHLBI) of the National Institutes of Health (NIH), a Cooperative Research and Development Agreement with Merck Sharpe & Dohme, and by funding from the trans-NIH Center for Human Immunology.

## Author Contributions

MG, GG, LWD, and CSH wrote the manuscript. MG, GG, and LWD made the figures. MG, LWD, KEL, JYG, KAO, YL, PKD, JLM, JD, JK, JLG, and CSH designed experiments. MG, LWD, KEL, and PKD performed experiments. MG, GG, LWD, DK, KAO, KRC, and CSH analyzed and interpreted data. JT, JV, KAO, CBD, TH, KRC, CL, and CSH were involved in the clinical care of the patients. DMS, HT, CL, and CHS developed the clinical concept and design CSH supervised the study. All authors reviewed and approved the final manuscript.

## Competing Interests

CSH receives research support from Merck Sharpe & Dohme and SELLAS Life Sciences Group AG. CL is on the Speakers’ Bureau for Astellas, Jazz Pharma and serves in a consulting or advisory role for Daiichi, Jazz Pharma, Amgen, Abbvie, Macrogenics, and Agios. The remaining authors declare no competing interests.

## Data and Materials Availability

Bulk TCRβ sequencing data is available on the ImmunoSEQ Analyzer through Adaptive Biotechnologies. scRNA-seq data sets are available through the National Center for Biotechnology Information’s Gene Expression Omnibus. Code available upon request.

## Supplementary Materials

**Fig. S1**. Frequency and diversity of BM-infiltrating and circulating T cells.

**Fig. S2**. Significantly differentially abundant T cell clones in patients developing irAEs.

**Fig. S3**. Comparable magnitude of expansion of clones of interest in BM and PB in 2 patients developing irAEs.

**Fig. S4**. Removal of doublets and low-quality cells in 5’ scRNA-seq data.

**Fig. S5**. Significantly expanded and novel clones detected at EOC4 by scRNA-seq in irAE timepoints.

**Fig. S6**. Downsampling of scRNA-seq data.

**Fig. S7**. Significantly differentially abundant T cell clones in patients with responsive disease.

**Fig. S8**. Significantly differentially abundant T cell clones in patients with progressive disease.

**Fig. S9**. Putative clones discovered by scRNA-seq in responders do not share irAE cell protein signature.

**Fig. S10**. Immunophenotyping of 5’ scRNA-seq data based on combinatorial cell surface protein expression.

**Fig. S11**. Comparison of T cell metrics between scRNA-seq, TCRβ-seq, and clinical flow cytometry.

**Fig. S12**. Phenotypes of pre-existing clones during treatment.

**Fig. S13**. Phenotyping of immune cells and leukemia in 3’v3 scRNA-seq and scDNA-seq.

**Table S1**. CD19+ B cell frequencies by IHC during treatment.

**Table S2**. Anti-thyroid autoantibody levels in patients developing hypothyroidism.

**Table S3**. QC of 5’scRNA-seq data with removal of cells with low CD3/CD45, and both low or both high CD4/CD8 expression.

**Table S4**. Eligibility assessment and enrollment criteria.

**Table S5**. Response and discontinuation criteria.

**Table S6**. Total Seq-C antibodies used with 5’ scRNA-seq.

**Table S7**. TotalSeq-A antibodies used with 3’v3 scRNA-seq.

## Notes

### Clinical Trial

NCT02996474

### Clinical Protocols

https://clinicaltrials.gov/ct2/show/NCT02996474

### Author Declarations

NHLBI IRB, protocol number 17-H-0026

