## Supplement for "Pembrolizumab and Decitabine for Refractory or Relapsed Acute Myeloid Leukemia"

FigS1

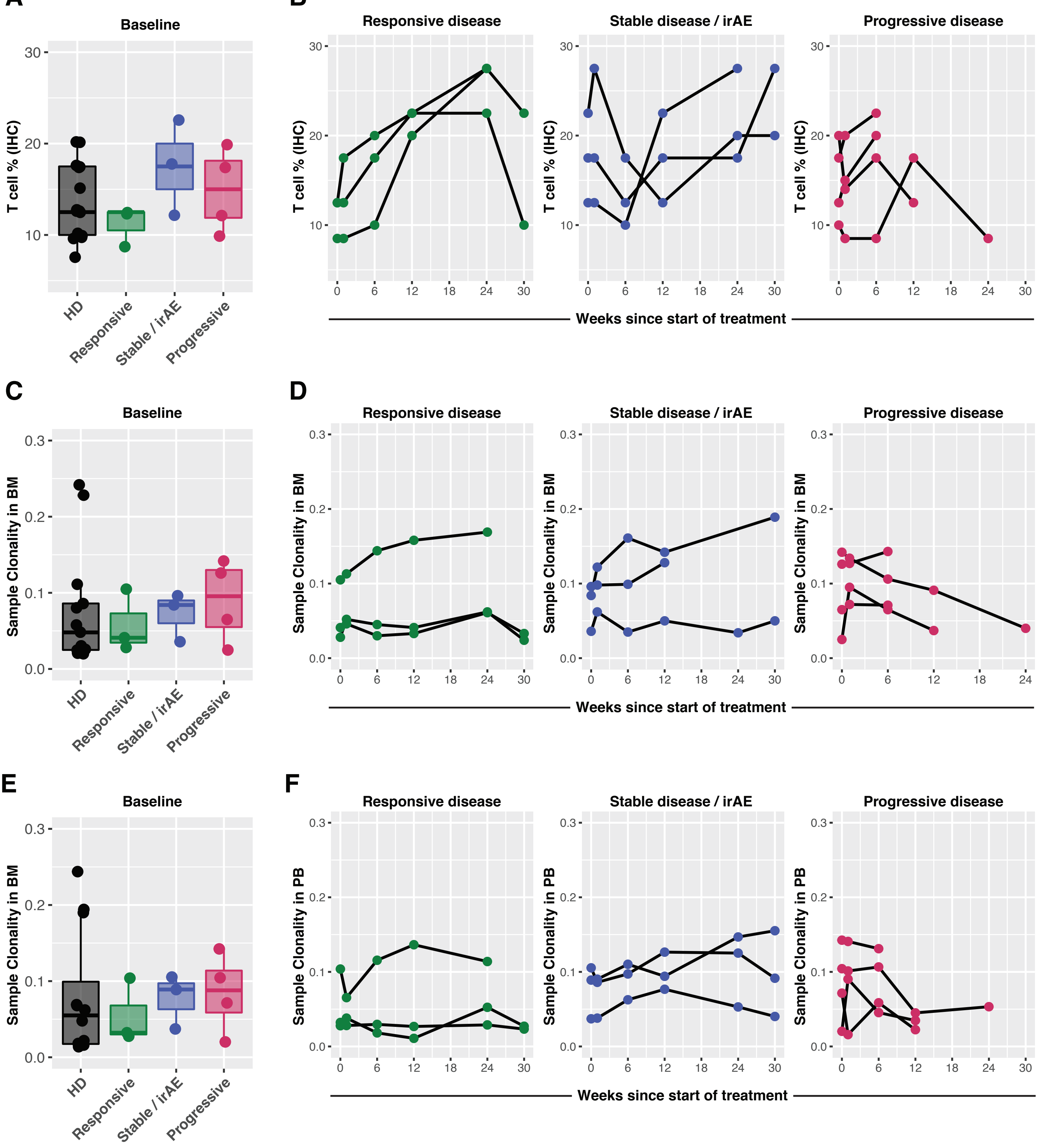


**Fig. S1. Frequency and diversity of BM-infiltrating and circulating T cells.** (A) Frequencies of CD3+ T cells at baseline in the BM of healthy donors (HD, n=13, previously reported in Williams P, Cancer 2019) and 10 enrolled patients, measured by immunohistochemistry (IHC) of BM core biopsies. (B) CD3+ T cell frequencies by IHC in BM of patients during treatment. BM TCRβ repertoire clonality in HD and PD-AML patients at (C) baseline and (D) during treatment. Circulating PB TCRβ repertoire clonality in HD and PD-AML patients at (E) baseline and (F) during treatment. For all panels, patients are grouped by response.

FigS2

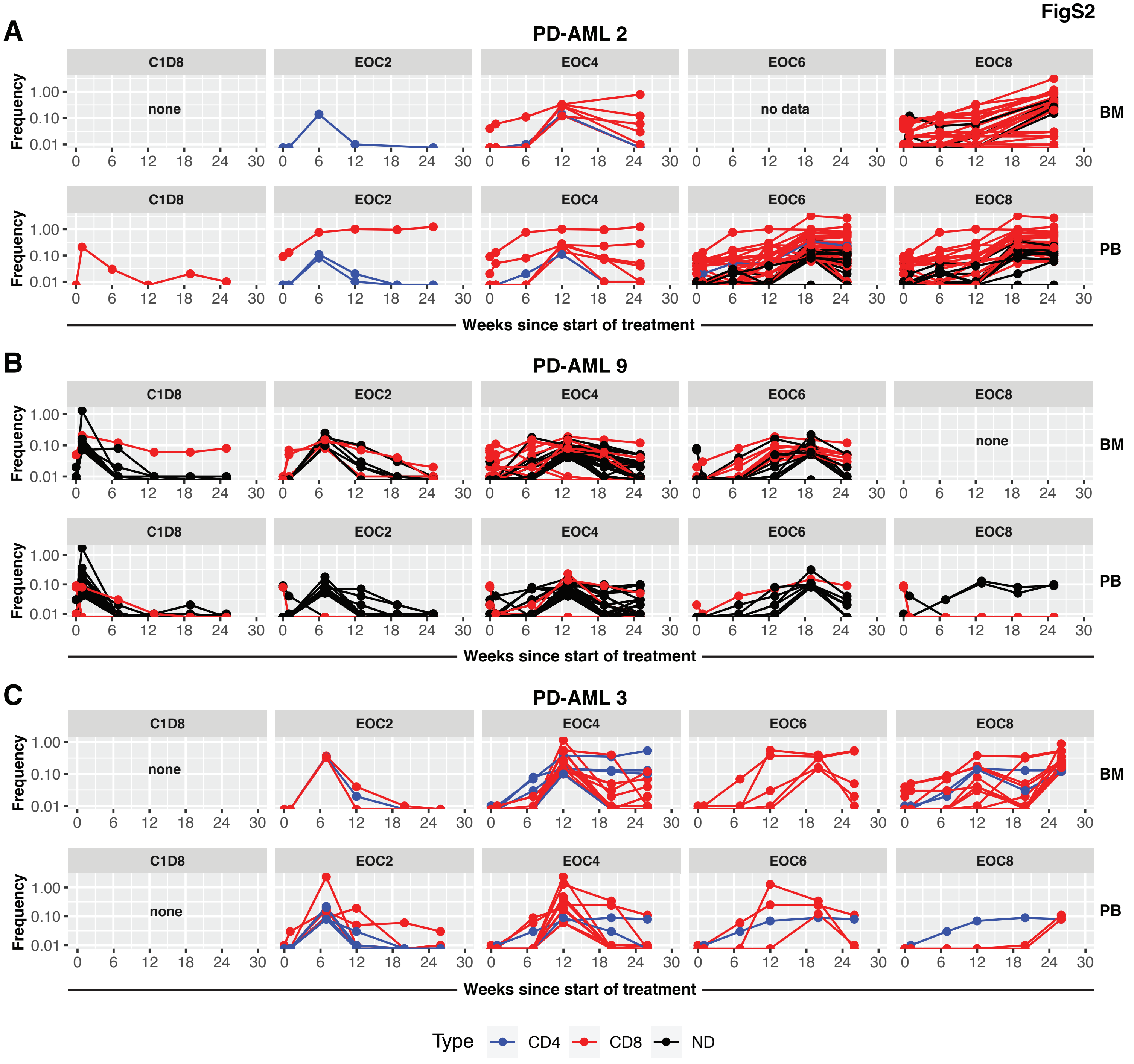


**Fig. S2. Significantly differentially abundant T cell clones in patients developing irAEs.** Pairwise identification of significantly abundant T cell clones between longitudinal timepoints compared to baseline for BM and PB in (A) PD-AML 2 (hypothyroidism at EOC4), (B) PD-AML 9 (hypothyroidism at EOC2), and (C) PD-AML 3 (central diabetes insipidus, at EOC4) was performed. Top and bottom row of each panel shows all significantly abundant clones with a baseline productive frequency of 0.1% or below in BM and PB, respectively. Red indicates CD8+ T cell clones; blue indicates CD8- (CD4+) T cell clones. Black indicates clones where CD4/CD8 could not be determined (ND).

FigS3

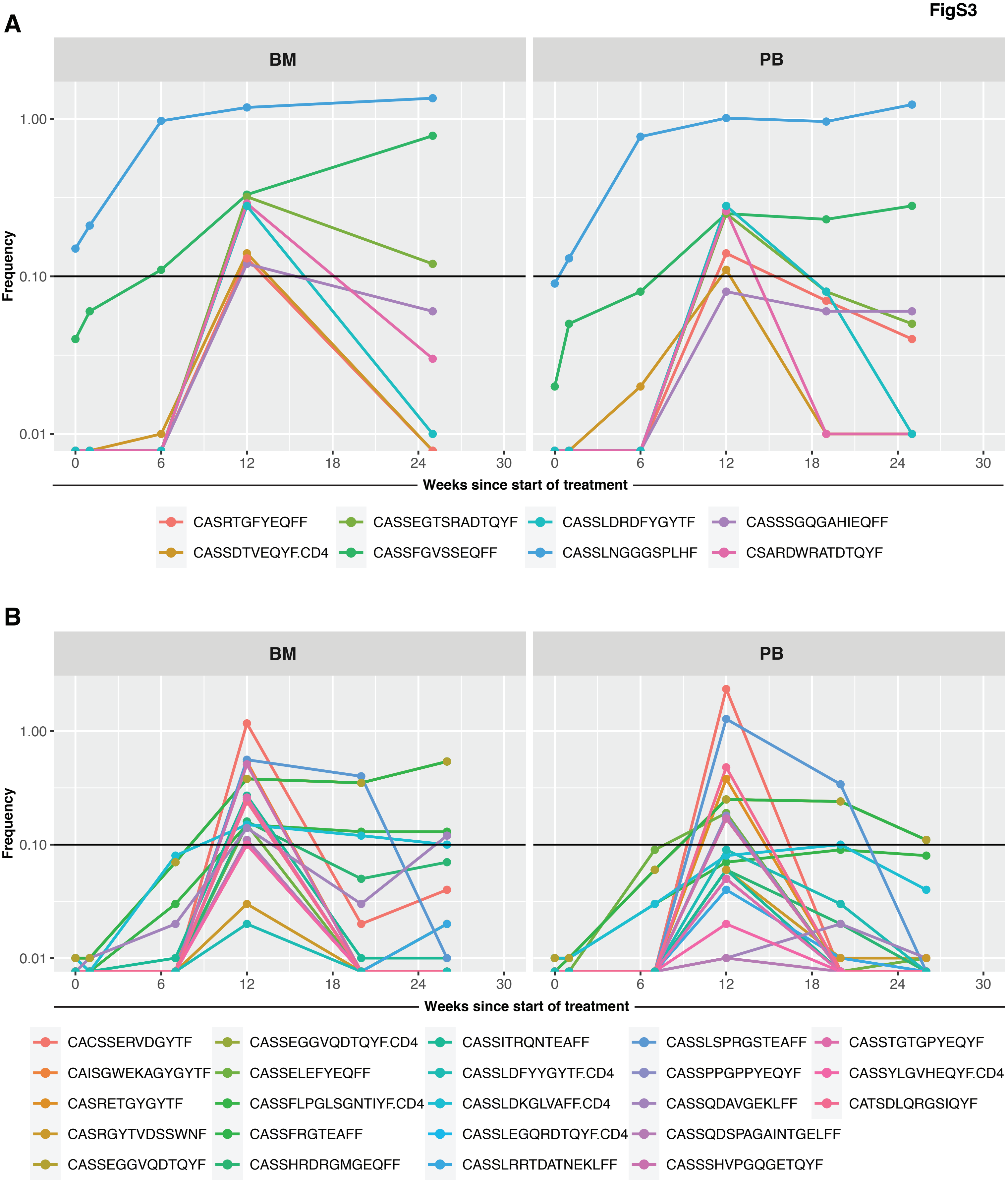


**Fig. S3. Comparable magnitude of expansion of clones of interest in BM and PB in 2 patients developing irAEs.** The frequencies of clones identified as significantly abundant at irAE onset that were low to undetectable in BM and/or PB in (A) PD-AML 2 (B) PD-AML 3. Time since start of treatment in weeks is indicated along x-axis for all plots.

FigS4

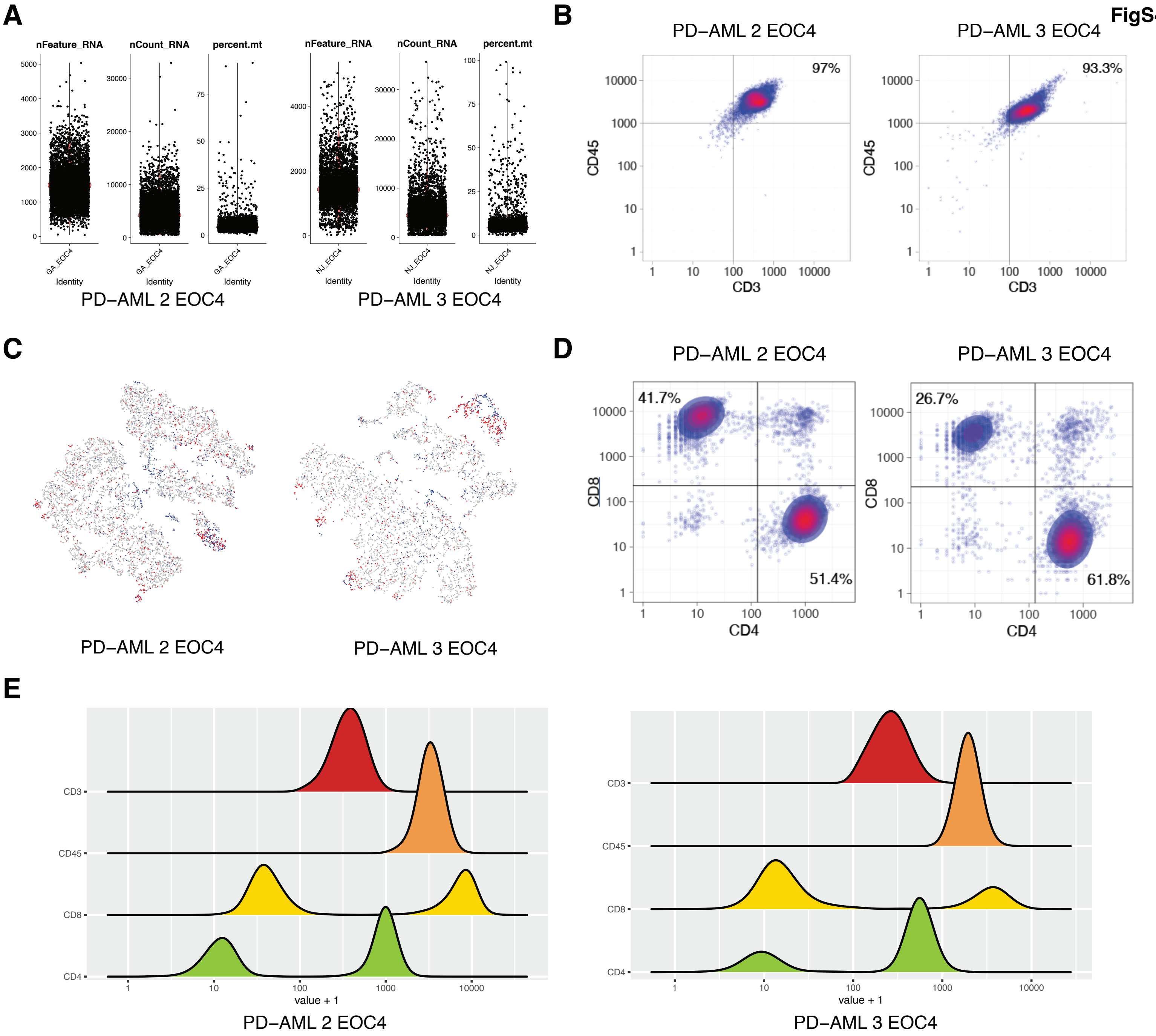


**Fig. S4. Removal of doublets and low-quality cells in 5' scRNA-seq data.** (A) Loose cutoffs for mitochondrial gene percentages and total gene expression level, including number of cell surface feature and total UMI counts, were set in initial filtering. (B) By observing the distribution of CD3 and CD45 expression, a global cutoff was determined for valid T cells (CD3 > 100, CD45 > 1000). (C) To further remove doublets, cells harboring 2 distinct TCRβ CDR3 sequences (highlighted in red) were removed. Cells in blue indicate cells that had no detected TCRβ CDR3; these cells were not removed. (D) To remove additional low-quality cells and double negative and double positive cells, the density of CD4 and CD8 expression was plotted and cells below or above both cutoffs (CD4: 130, CD8: 228) were excluded. (E) Cutoffs for all markers were further confirmed by looking at the distributions of markers on ridge plots and by model-based clustering. Data for PD-AML 2 and PD-AML 3 at EOC4 are shown in all panels.

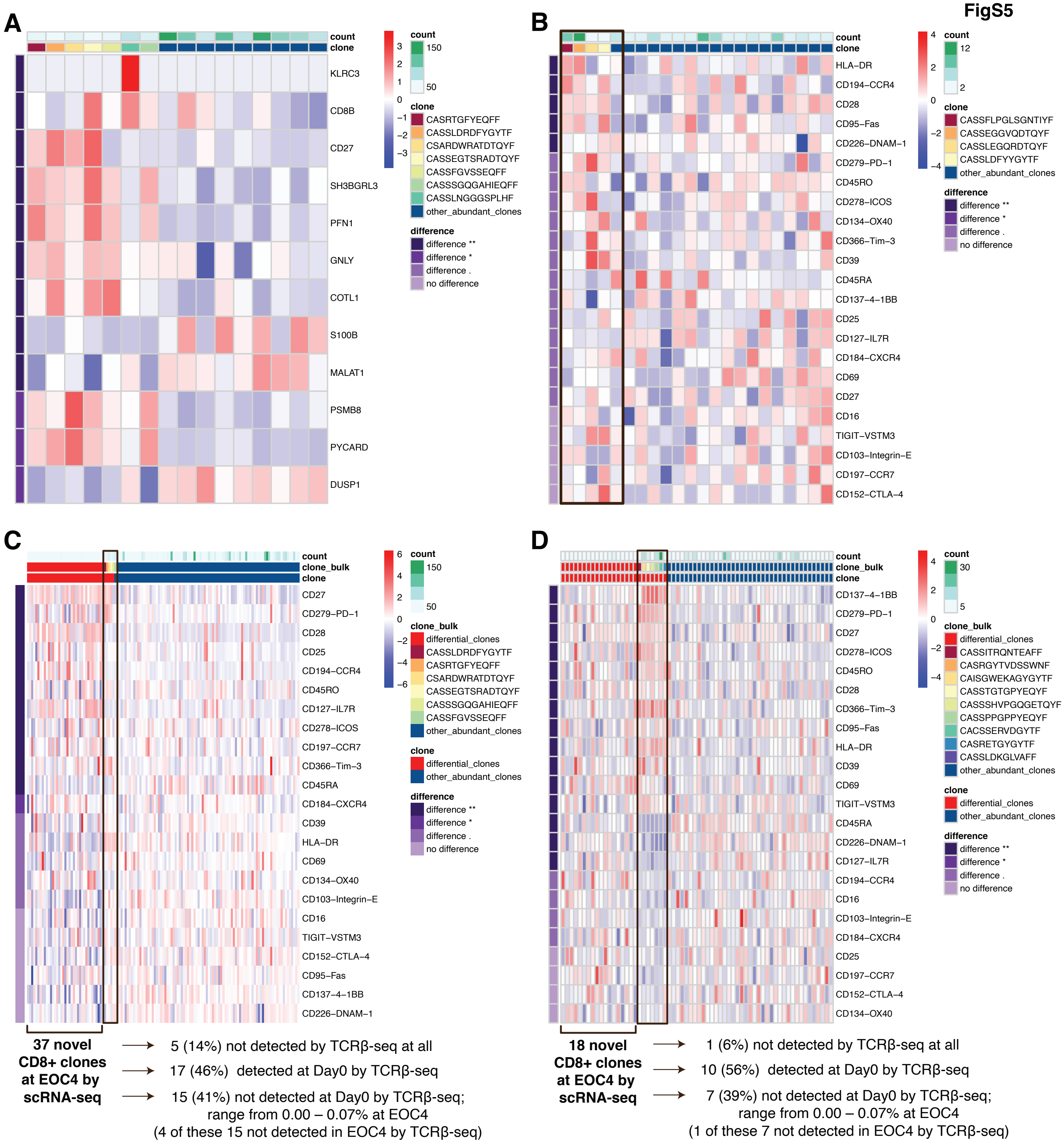


**Fig. S5. Significantly expanded and novel clones detected at EOC4 by scRNA-seq in irAE timepoints.** (A) Differential gene expression profile of significantly expanded clone CASSLNGGGSPLHF in PD-AML 2 EOC4. (B) Cell surface expression profile of significantly expanded CD4+ clones of interest in PD-AML 3 at EOC4. (C-D) Differential cell surface protein expression of novel T cell clones by scRNA-seq in PD-AML 2 at EOC4 and at PD-AML 3 at EOC4, respectively, that were not detected at Day0 or EOC2 timepoints by scRNA-seq. Clones of interest that were identified by bulk TCRβ-seq are grouped together (indicated by box), and the absence or presence in the bulk TCRβ-seq data of remaining novel clones by scRNA-seq is described. Individual T cell clones, expression scale, cells per clone, and significance level are indicated next to each heat map.

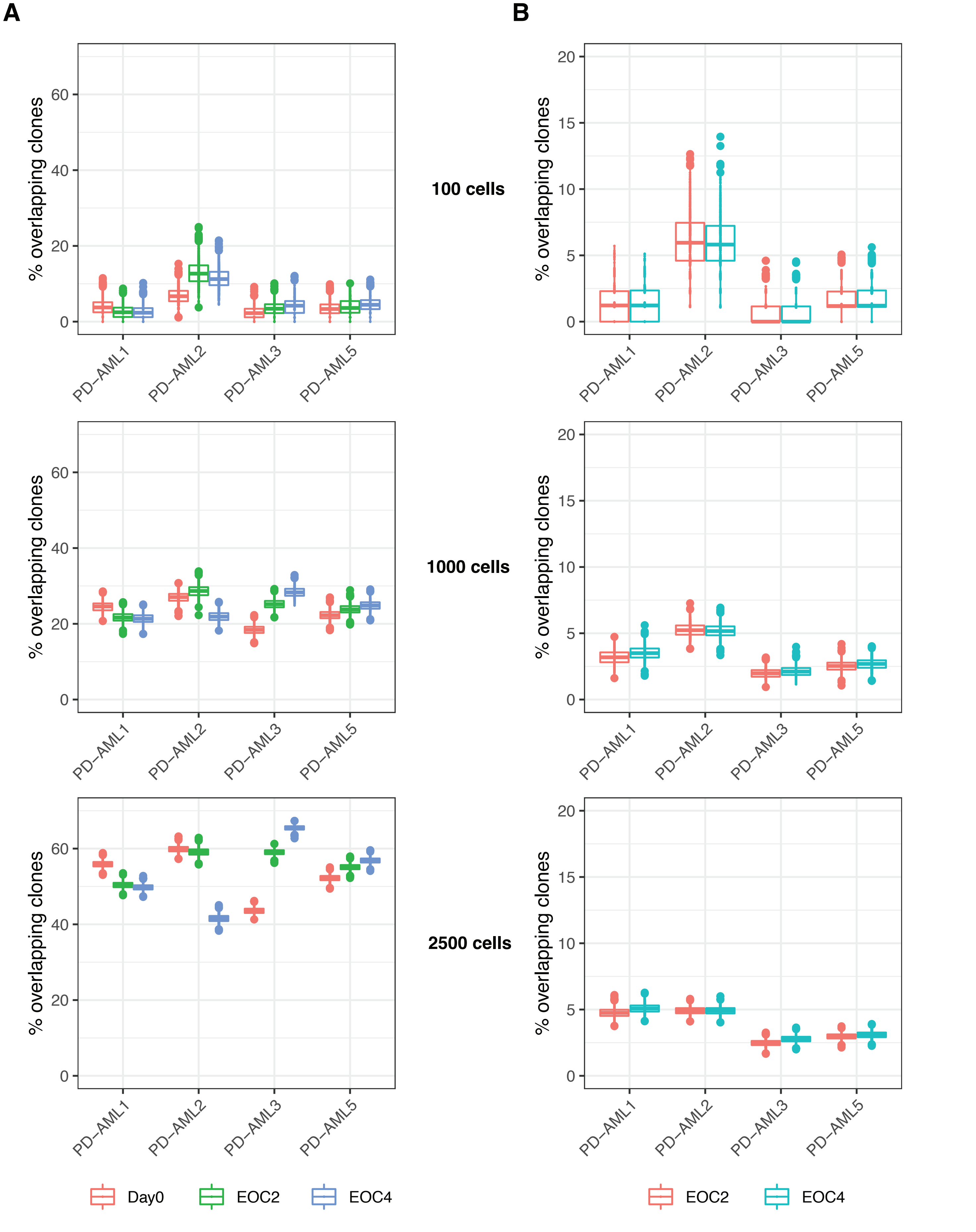


**Fig. S6. Downsampling of scRNA-seq data.** (A) T cell scRNA-seq datasets were downsampled without replacement to 100, 1000, and 2500 cells to compare overlap in shared and unique clones. The percentages of overlapping clones between 2 equal-sized samplings within individual patient timepoints were calculated and the approximate distribution of 1000 downsampling iterations was plotted. Patient sample timepoints are indicated in the legend. (B) Samples were downsampled without replacement to 100, 1000, and 2500 cells, and the percentage of overlapping clones between EOC2 or EOC4 with Day0 was calculated for 1000 iterations. Patient sample timepoints indicated in the legend.

FigS7

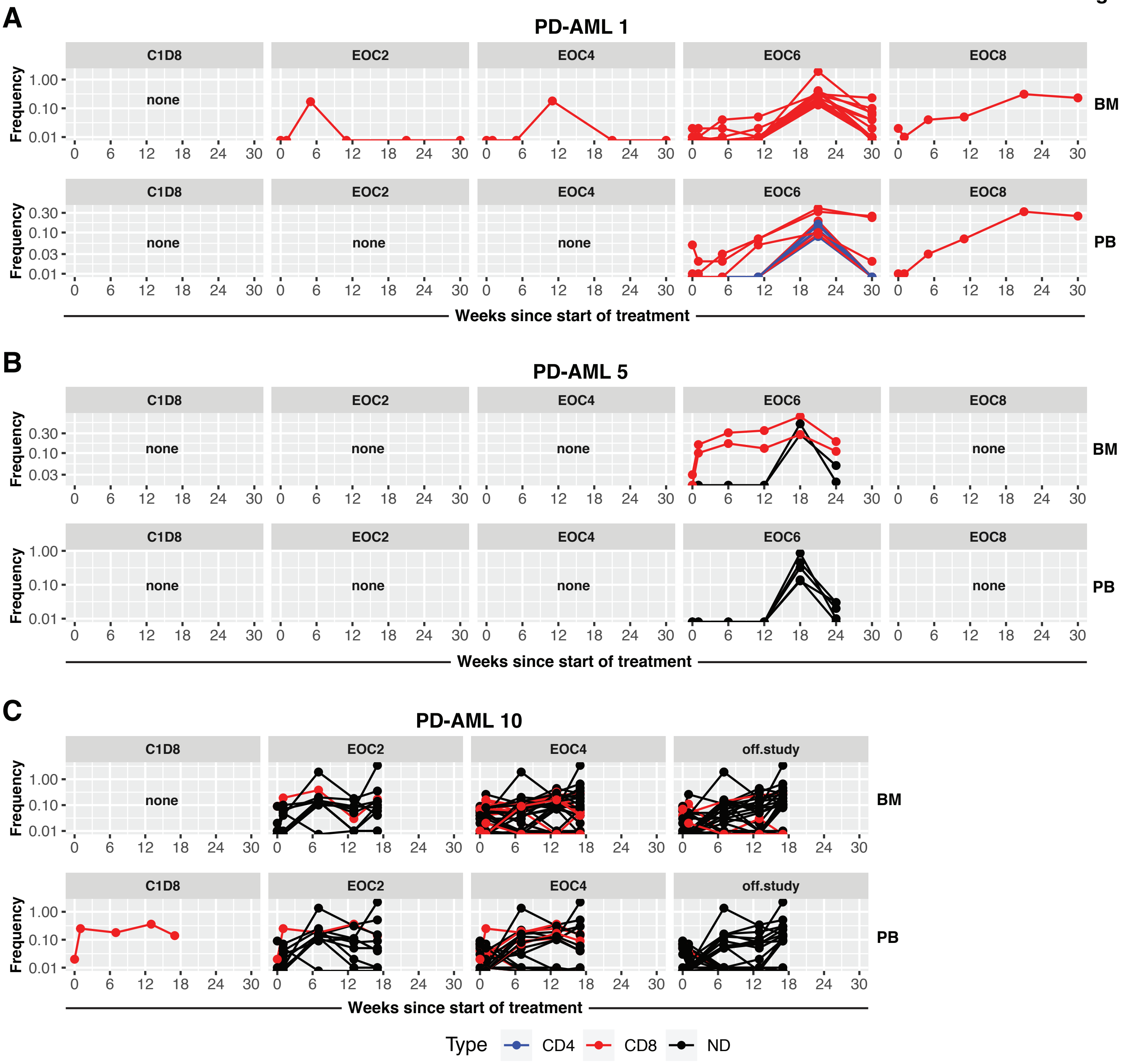


**Fig. S7. Significantly differentially abundant T cell clones in patients with responsive disease.** Pairwise identification of significantly abundant T cell clones between longitudinal timepoints compared to baseline for BM and PB in (A) PD-AML 1, (B) PD-AML 5, and (C) PD-AML 10 (taken off study mid-cycle 5) was performed. Top and bottom row of each panel shows all significantly abundant clones with a baseline productive frequency of 0.1% or below in BM and PB, respectively. Red indicates CD8+ T cell clones; blue indicates CD8- (CD4+) T cell clones. Black indicates clones where CD4/CD8 could not be determined (ND).

FigS8

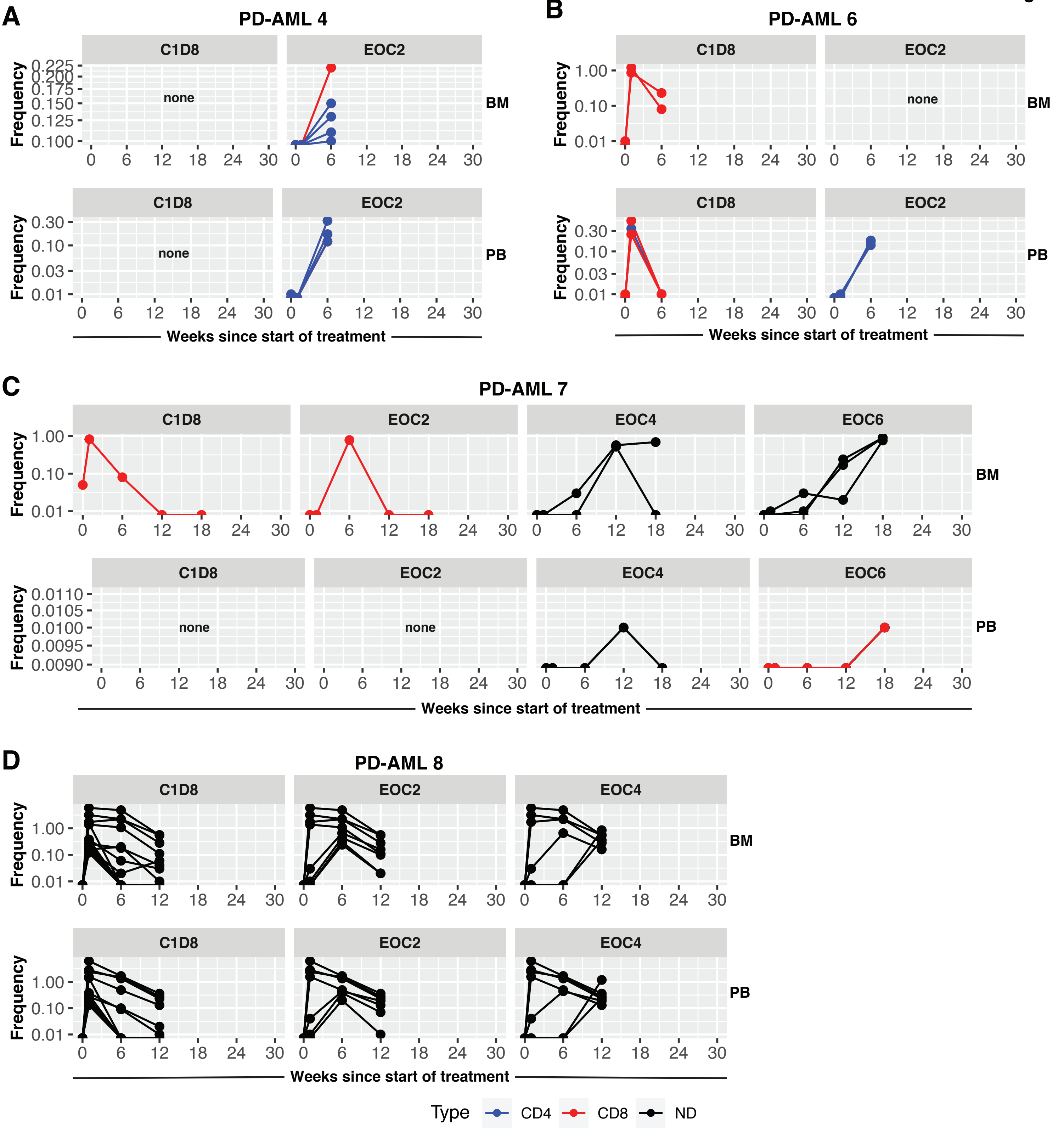


**Fig. S8. Significantly differentially abundant T cell clones in patients with progressive disease**. Pairwise identification of significantly abundant T cell clones between longitudinal timepoints compared to baseline for BM and PB in (A) PD-AML 4, (B) PD-AML 6, (C) PD-AML 7, and (D) PD-AML 8 was performed as described in Fig. S2. Top and bottom row of each panel shows all significantly abundant clones with a baseline productive frequency of 0.1% or below in BM and PB, respectively. Red indicates CD8+ T cell clones; blue indicates CD8- (CD4+) T cell clones. Black indicates clones where CD4/CD8 could not be determined.

FigS9

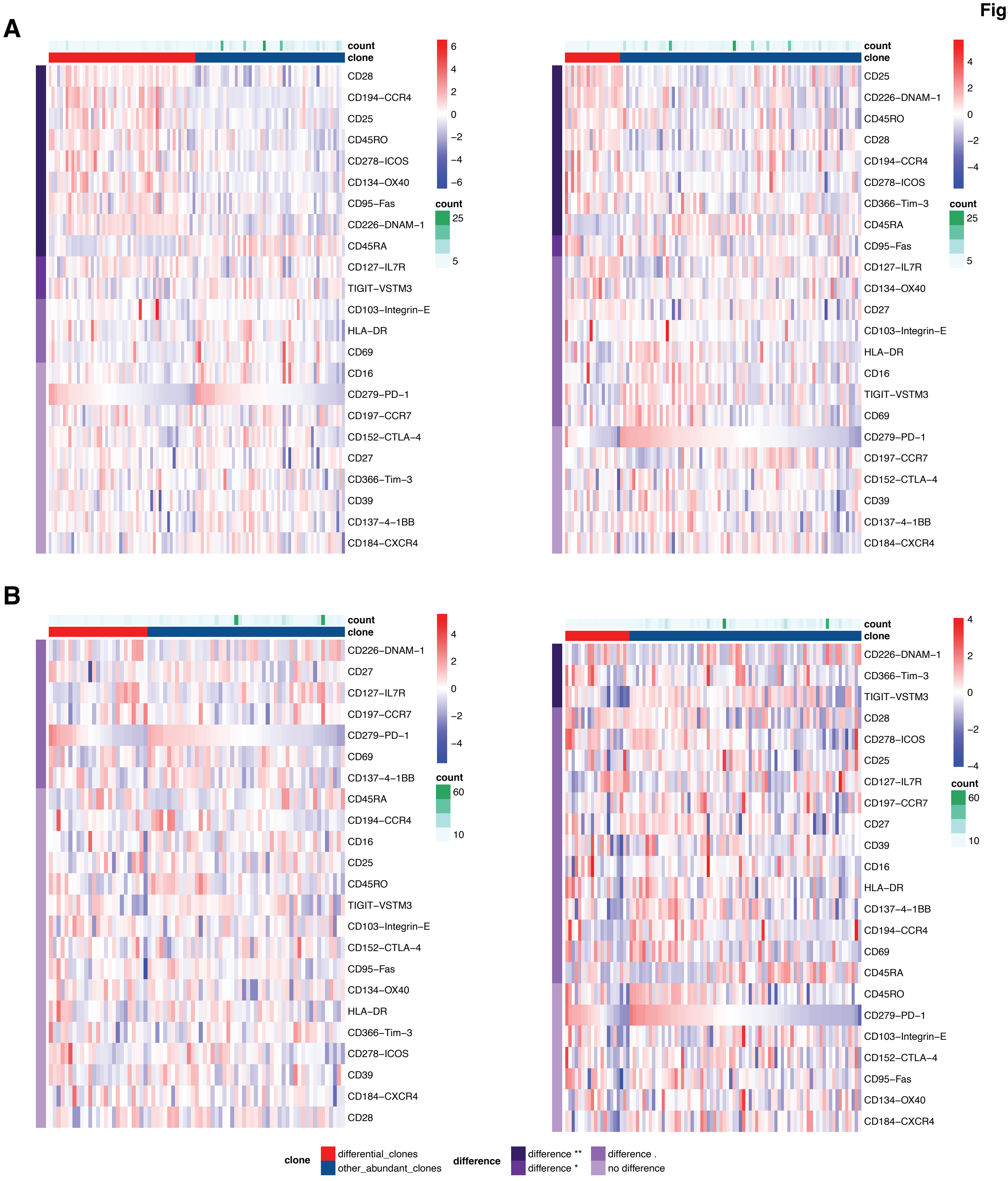


**Fig. S9. Putative CD8+ T cell clones discovered by scRNA-seq in responders do not share irAE cell protein signature.** Supervised analysis of DEP between clones detected with scRNA-seq in (A) PD-AML 1 at EOC2 (left) and EOC4 (right) and (B) PD-AML 5 at EOC2 (left) and EOC4 (right) that were not found in the baseline sample of the respective patient. Heatmaps were created using the mean expression of all cells within each clone, and clonotypes are ranked based on relative PD-1 expression. Differentially expressed markers were ordered by adjusted p-values and fold changes. Expression scale and level of difference are indicated by each heat map.

FigS10

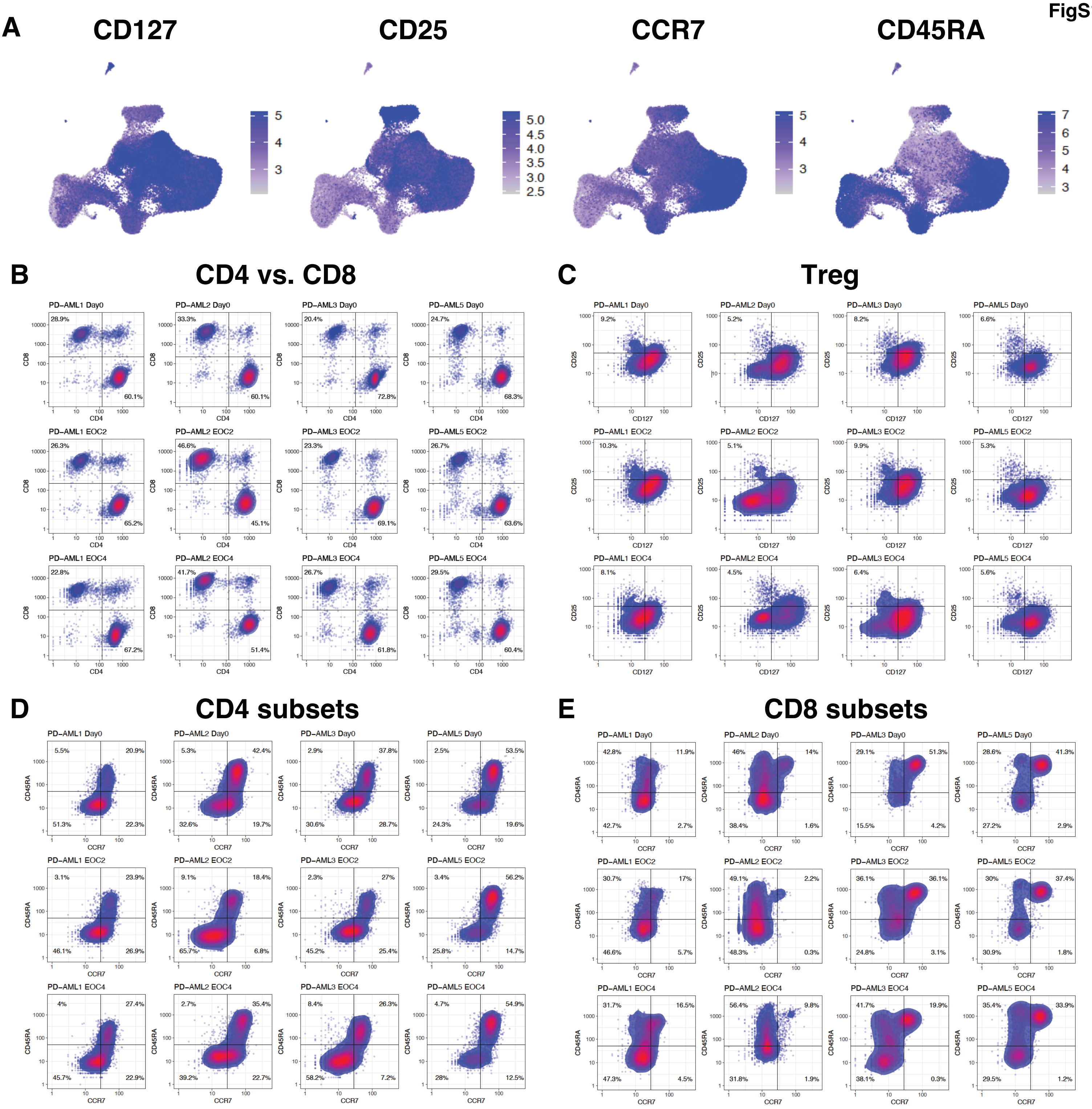


**Fig. S10. Immunophenotyping of 5’ scRNA-seq data based on combinatorial cell surface protein expression.** (A) Expression of CD3, CD45, CD4, and CD8 were used in quality control as described in Fig. S4; these markers in addition to CD127, CD25, CCR7, and CD45RA (purple) were used for T cell subset immunophenotyping. Data from all patients and timepoints is visualized. Consensus cutoffs were created by comparing median cutoffs across different timepoints within one sample and adjusting them across all patients. (B) Criteria described in Fig. S4 were applied to remove low-quality, double negative, and double positive T cells based on CD4 and CD8 expression. (C) Tregs were identified based on high CD25 (cutoff >51) and low CD127 (cutoff <26) expression. Naïve (CCR7+CD45RA+), central memory (CM, CCR7+CD45RA-), effector memory (EM,CCR7-CD45RA-), and terminal effector cells (TE) that have re-expressed CD45RA (CCR7-CD45RA+) were gated based on CCR7 (cutoff: 28) and CD45RA (cutoff: 51) expression

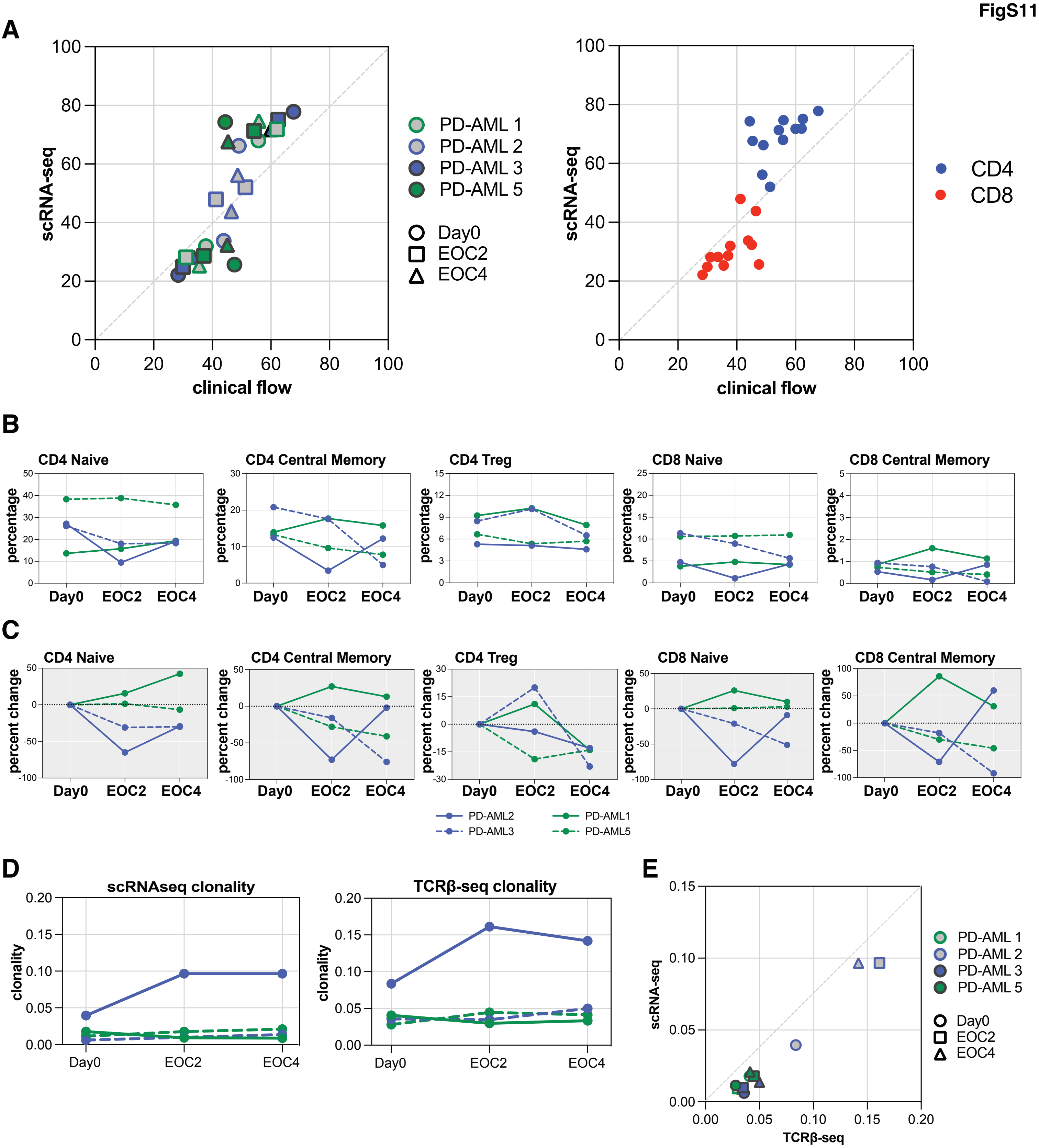


**Fig. S11. Comparison of T cell metrics between scRNA-seq, TCRβ-seq, and clinical flow cytometry.** (A) Correlation of CD4+ and CD8+ T cell frequencies as percent of CD3+ as calculated by scRNA-seq and clinical flow cytometry in patients across timepoints (left), with CD4 or CD8 T cell compartment specified (right). (B) Frequencies of CD4+ and CD8+ naïve and central memory (CM) T cells and Tregs at Day0, EOC2, and EOC4, as a percentage of CD3 at each timepoint. (C) Percent change in frequencies of above T cell subpopulations at EOC2 and EOC4 compared to Day0 percentage. (D) Bulk clonality measures calculated from scRNA-seq (left) and TCRβ-seq (right). Patients indicated by legend. (E) Comparison of clonality measures for each sample calculated by both modalities. Patients and timepoints indicated in legend.

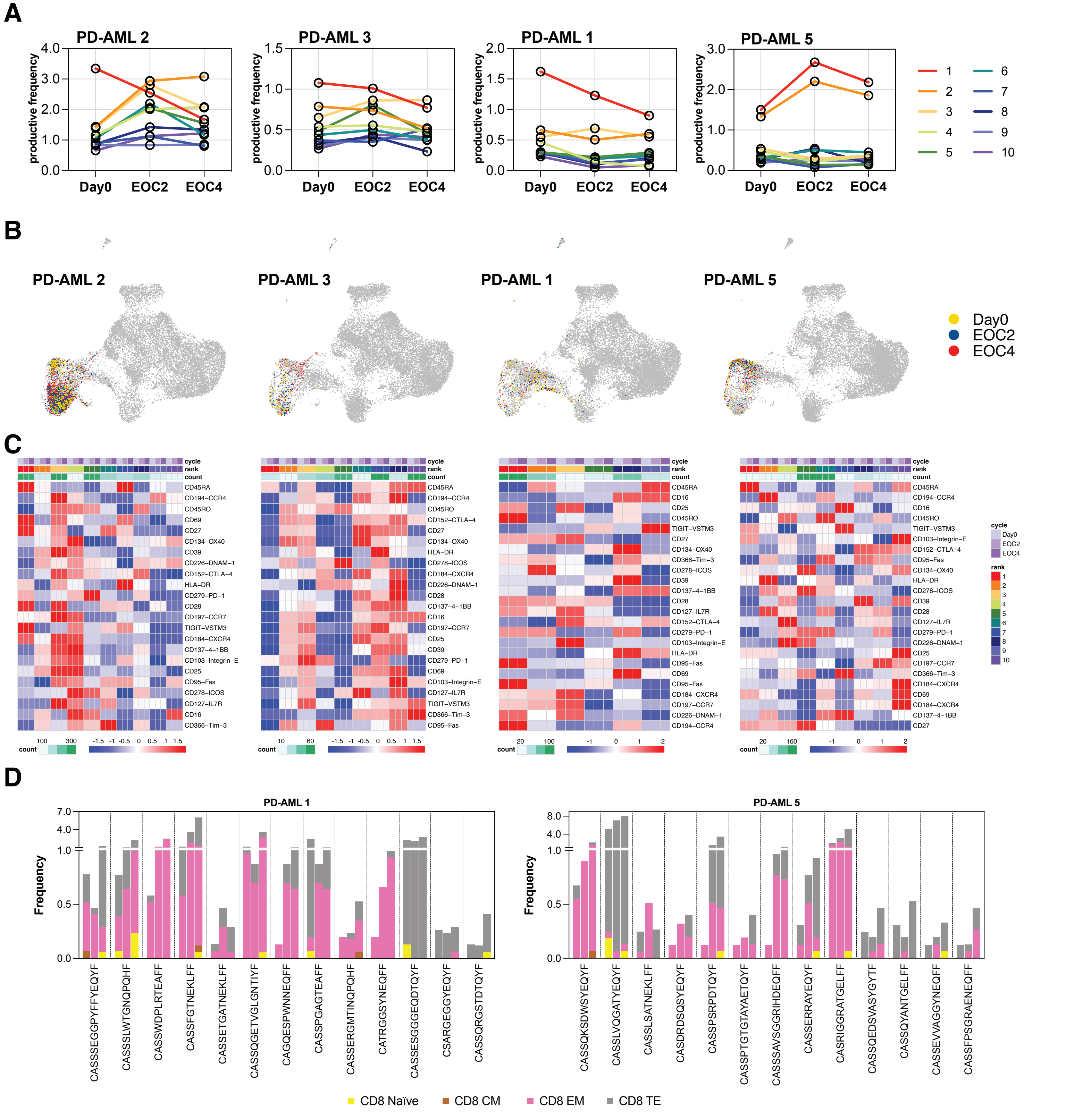


**Fig. S12. Phenotypes of pre-existing clones during treatment.** (A) Frequencies at Day0, EOC2, and EOC4 of the top 10 most abundant clonotypes at baseline from bulk TCRβ-seq. Clones are color coded by rank indicated in legend. (B) Visualization of top 10 abundant clones from bulk TCRβ-seq in scRNA-seq integrated UMAP. (C) Cells containing the same TCRβ identified from bulk TCRβ-seq were selected in the scRNA-seq data if the clones were present at all sampled timepoints. Average expression of cell surface proteins from each clone was plotted using normalized count data within each sample. Expression scale, timepoint, and abundant clone rank is indicated by the legend, and the number of detected cells per clone is summarized by the scale below each heat map. (D) Clonotypes were selected from scRNA-seq data from responders if the clonotype was comprised of more than one phenotype (naïve, CM, EM, or TE). The frequency of each phenotype per clonotype is plotted with first bar indicating Day0, second bar EOC2, and third bar EOC4. T cell phenotypes indicated in the legend.

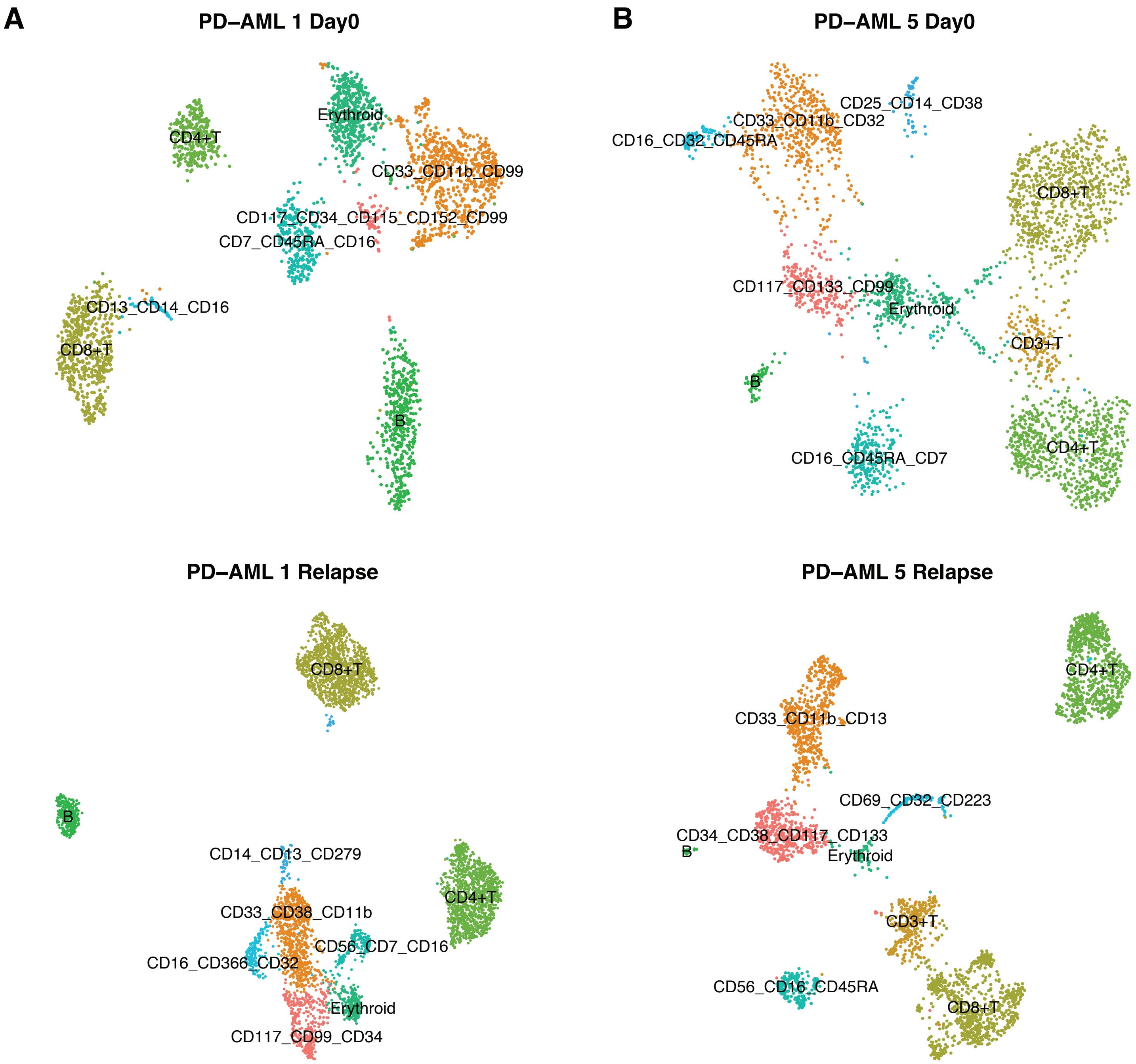


**Fig. S13. Phenotyping of immune cells and leukemia in 3'v3 scRNA-seq and scDNA-seq.** Clustering based on cell surface protein expression of 3'v3 scRNA-seq acquired from BMMC was performed for (A) PD-AML 1 Day0 (top) and relapse (bottom) and (B) PD-AML 5 Day0 (top) and relapse (bottom). Clusters highly expressing CD3 and CD4/CD8 were labeled as T cells, and those with high CD19 and CD22 expression were annotated as B cells. Erythroid cells were identified as cells with no highly expressed proteins and confirmed by hemoglobin gene expression. For myeloid lineages, clusters with high expression of CD45RA/CD7 and CD16 were considered healthy monocytes. Putative leukemic cells were identified based on expression of known leukemia-associated markers for each patient.

**Table S1. CD19+ B cell frequencies by IHC during treatment.**

| Patient | Day0 | C1D8 | EOC2 | EOC4 | EOC6 | EOC8 |
|---|---|---|---|---|---|---|
| PD-AML 1 | 5 | 5 | 3 | 3 | 2 | 2 |
| PD-AML 2 | < 1 | < 1 | < 1 | < 1 | - | - |
| PD-AML 3 | < 1 | < 1 | < 1 | < 1 | < 1 | < 1 |
| PD-AML 4 | < 1 | 1.5 | < 1 | - | - | - |
| PD-AML 5 | 1.5 | 2.5 | 1.5 | 1.5 | < 1 | < 1 |
| PD-AML 6 | 2 | 1 | 1 | - | - | - |
| PD-AML 7 | 3 | 2 | 1.5 | 1.5 | 1 | - |
| PD-AML 8 | 4 | 4 | 5 | 4 | - | - |
| PD-AML 9 | < 1 | < 1 | < 1 | < 1 | < 1 | < 1 |
| PD-AML 10 | < 1 | < 1 | < 1 | < 1 | < 1 * | - |

- denotes unavailable data; * indicates mid-cycle off-study timepoint for this patient

**Table S2. Anti-thyroid autoantibody levels in patients developing hypothyroidism.**

| Patient | Thyroid hormone levels at irAE onset | | Baseline antibodies (IU/mL) | | Antibodies at irAE onset (IU/mL) | |
|---|---|---|---|---|---|---|
| | TSH | Free T4 | anti-TG | anti-TPO | anti-TG | anti-TPO |
| PD-AML 2 | 67 | 0.4 | <20.0 | <10.0 | 21.6 | 19.8 |
| PD-AML 9 | 22 | - | - | - | <20.0 | <10.0 |
| Reference Range | 0.27-4.2 mcIU/mL | 0.9-1.7 ng/dL | 0.0-40.0 IU/mL | 0.0-34.9 IU/mL | 0.0-40.0 IU/mL | 0.0-34.9 IU/mL |

- denotes unavailable data; values in red outside normal ranges; TSH, thyroid stimulationg hormone; T4, thyroxin; TG, thyroglobulin; TPO, thyroid peroxidase

**Table S3. QC of 5'scRNA-seq data with removal of cells with low CD3/CD45, and both low or both high CD4/CD8 expression.**

| Patient | Timepoint | Acquired Cell Number | RNA count* | >1 CDR3** | CD3/CD45 † | CD4/CD8 ‡ | Final Cell Number |
|---|---|---|---|---|---|---|---|
| PD-AML 2 | Day0 | 5480 | 258 | 437 | 263 | 184 | 4338 |
| | EOC2 | 6011 | 264 | 574 | 450 | 266 | 4457 |
| | EOC4 | 8612 | 457 | 735 | 241 | 363 | 6816 |
| PD-AML 3 | Day0 | 7193 | 286 | 612 | 154 | 239 | 5902 |
| | EOC2 | 5245 | 81 | 435 | 153 | 235 | 4341 |
| | EOC4 | 5519 | 644 | 466 | 302 | 225 | 3882 |
| PD-AML 1 | Day0 | 6084 | 363 | 460 | 258 | 365 | 4638 |
| | EOC2 | 6548 | 275 | 356 | 391 | 372 | 5154 |
| | EOC4 | 6842 | 426 | 547 | 370 | 358 | 5141 |
| PD-AML 5 | Day0 | 6071 | 310 | 355 | 308 | 205 | 4893 |
| | EOC2 | 6006 | 285 | 437 | 442 | 182 | 4660 |
| | EOC4 | 5695 | 239 | 395 | 309 | 231 | 4521 |

* removal based on mitchondrial gene content and UMI counts; ** removal of cells containing more than 1 TRB gene; † removal of cells without cell surface expression of CD3 and CD45; ‡ removal of cells expressing both or no cell surface CD4 and CD8

| Table S4. Eligibility assessment and enrollment criteria. | |
|---|---|
| | • Unequivocal diagnosis of relapsed or refractory acute myeloid leukemia (AML) confirmed by an NIH attending pathologist within 30 days of study enrollment (includes residual AML as confirmed by institutional standards by NIH pathologists. |
| | • Received at least one prior AML therapy before study enrollment. |
| | • Ability to comprehend the investigational nature of the study and provide informed consent. |
| | • Be at least 18 years of age on day of signing informed consent. |
| | • Availability of a physician willing to assume clinical care after completion of the study. |
| | • Be willing to provide blood and bone marrow for research as described in the study. |
| | • Have a performance status of less than or equal to 2 on the ECOG Performance Scale. |
| ***Inclusion Criteria*** | • Demonstrate adequate organ function; all screening labs should be performed within 14 days of treatment initiation. |
| | • Female subject of childbearing potential should have a negative urine or serum pregnancy within 72 hours prior to receiving the first dose of study medication. If the urine test is positive or cannot be confirmed as negative, a serum pregnancy test will be required. |
| | • Female subjects of childbearing potential must be willing to use an adequate method of contraception for the course of the study through 120 days after the last dose of study medication. Note: Abstinence is acceptable if this is the usual lifestyle and preferred contraception for the subject. |
| | • Male subjects of childbearing potential must agree to use an adequate method of contraception, starting with the first dose of study therapy through 120 days after the last dose of study therapy. Note: Abstinence is acceptable if this is the usual lifestyle and preferred contraception for the subject. |
| | • Has a diagnosis of acute promyelocytic leukemia (APL) |
| | • Has previously received an allogeneic hematopoietic stem cell transplant.f acute promyelocytic leukemia (APL). |
| | • Has received AML treatment with an investigational therapy or device within 4 weeks of the first dose of treatment. |
| | • Has a diagnosis of immunodeficiency or is receiving systemic steroid therapy or any other form of immunosuppressive therapy within 7 days prior to the first dose of trial treatment. |
| | • Has a known history of active TB (Bacillus Tuberculosis) |
| | • Has hypersensitivity to pembrolizumab or any of its excipients. |
| | • Has hypersensitivity to decitabine or any of its excipients. |
| | • Has received more than two prior cycles of decitabine. |
| | • Has had a prior anti-cancer monoclonal antibody (mAb) within 4 weeks prior to study Day 1. |
| | • Has had prior chemotherapy, targeted small molecule therapy, or radiation therapy within 2 weeks prior to study Day 1. Subjects who have received cytoreductive therapy with hydroxyurea at any time prior to study Day 1 are an exception to this criterion. |
| | • Has not recovered (i.e., ≤ Grade 1 or at baseline) from adverse events due to previously administered AML therapy agents. Subjects with ≤ Grade 2 neuropathy are an exception and may qualify for the study. If subject received major surgery, they must have recovered adequately from the toxicity and/or complications from the intervention prior to starting therapy. |
| ***Exclusion Criteria*** | • Has a known additional malignancy that is progressing or requires active treatment e.g.: basal cell carcinoma of the skin or squamous cell carcinoma of the skin that has undergone potentially curative therapy or in situ cervical cancer. |
| | • Has known malignant central nervous system (CNS) involvement. |
| | • Has active autoimmune disease that has required systemic treatment in the past 2 years (i.e. with use of disease modifying agents, corticosteroids or immunosuppressive drugs). Replacement therapy (eg., thyroxine, insulin, or physiologic corticosteroid replacement therapy for adrenal or pituitary insufficiency, etc.) is not considered a form of systemic treatment. |
| | • Has history of (non-infectious) pneumonitis that required steroids, evidence of interstitial lung disease or active, non-infectious pneumonitis. |
| | • Has an uncontrolled active infection. |
| | • Has a history or current evidence of any condition, therapy, or laboratory abnormality that might confound the results of the trial, interfere with the subject's participation for the full duration of the trial, or is not in the best interest of the subject to participate, in the opinion of the treating investigator. |
| | • Has known psychiatric or substance abuse disorders that would interfere with cooperation with the requirements of the trial. |
| | • Is pregnant or breastfeeding, or expecting to conceive or father children within the projected duration of the trial, starting with the pre-screening or screening visit through 120 days after the last dose of trial treatment. |
| | • Has received prior therapy with an anti-PD-1, anti-PD-L1, or anti-PD-L2 agent. |
| | • Has a known history of Human Immunodeficiency Virus (HIV) (HIV 1/2 antibodies). |
| | • Has known active Hepatitis B (e.g., HBsAg reactive) or Hepatitis C (e.g., HCV RNA [qualitative] is detected). |
| | • Has received a live vaccine within 30 days of planned start of study therapy. Note: Seasonal influenza vaccines for injection are generally inactivated flu vaccines and are allowed; however intranasal influenza vaccines (e.g., Flu-Mist®) are live attenuated vaccines, and are not allowed. |

**Table S5. Response and discontinuation criteria.**

| Toxicity | Hold for Grade | Timing for Restarting Treatment | Treatment Discontinuation |
|---|---|---|---|
| Diarrhea / colitis | 2-3 | Toxicity resolves to Grade 0-1 | Toxicity does not resolve within 12 weeks of last dose or inability to reduce corticosteroid to 10 mg or less of prednisone or equivalent per day within 12 weeks |
| | 4 | Permanently discontinue | Permanently discontinue |
| AST, ALT, or Increased Bilirubin | 2 | Toxicity resolves to Grade 0-1 | Toxicity does not resolve within 12 weeks of last dose |
| | 3-4 | Permanently discontinue (see exception below)* | Permanently discontinue |
| Type 1 diabetes mellitus (if new onset) or Hyperglycemia | T1DM or 3-4 | Hold pembrolizumab for new onset Type 1 diabetes mellitus or Grade 3-4 hyperglycemia associated with evidence of beta cell failure | Resume pembrolizumab when patients are clinically and metabolically stable |
| Hypophysitis | 2-4 | Toxicity resolves to Grade 0-1. Therapy with pembrolizumab can be continued while endocrine replacement therapy is instituted | Toxicity does not resolve within 12 weeks of last dose or inability to reduce corticosteroid to 10 mg or less of prednisone or equivalent per day within 12 weeks |
| Hyperthyroidism | 3 | Toxicity resolves to Grade 0-1 | Toxicity does not resolve within 12 weeks of last dose or inability to reduce corticosteroid to 10 mg or less of prednisone or equivalent per day within 12 weeks |
| | 4 | Permanently discontinue | Permanently discontinue |
| Hypothyroidism | | Therapy with pembrolizumab can be continued while thyroid replacement therapy is instituted | Therapy with pembrolizumab can be continued while thyroid replacement therapy is instituted |
| Infusion Reaction | 2* | Toxicity resolves to Grade 0-1 | Permanently discontinue if toxicity develops despite adequate premedication |
| | 3-4 | Permanently discontinue | Permanently discontinue |
| Pneumonitis | 2 | Toxicity resolves to Grade 0-1 | Toxicity does not resolve within 12 weeks of last dose or inability to reduce corticosteroid to 10 mg or less of prednisone or equivalent per day within 12 weeks |
| | 3-4 | Permanently discontinue | Permanently discontinue |
| Renal Failure or Nephritis | 2 | Toxicity resolves to Grade 0-1 | Toxicity does not resolve within 12 weeks of last dose or inability to reduce corticosteroid to 10 mg or less of prednisone or equivalent per day within 12 weeks |
| | 3-4 | Permanently discontinue | Permanently discontinue |
| All Other Drug- Related Toxicity** | 2 | Toxicity resolves to Grade 0-1 | Toxicity does not resolve within 12 weeks of last dose or inability to reduce corticosteroid to 10 mg or less of prednisone or equivalent per day within 12 weeks |
| | 3-4 | Permanently discontinue | Permanently discontinue |

* If symptoms resolve within one hour of stopping drug infusion, the infusion may be restarted at 50% of the original infusion rate (e.g., from 100 mL/hr to 50 mL/hr). Otherwise dosing will be held until symptoms resolve and the subject should be premedicated for the next scheduled dose. '** Patients with intolerable or persistent Grade 2 drug-related AE may hold study medication at physician discretion. Permanently discontinue study drug for persistent Grade 2 adverse reactions for which treatment with study drug has been held, that do not recover to Grade 0-1 within 12 weeks of the last dose.

**Table S6. Total Seq-C antibodies used with 5' scRNA-seq.**

| Specificity | Clone | Barcode Sequence |
|---|---|---|
| CD3 | UCHT1 | CTCATTGTAACTCCT |
| CD8 | SK1 | GCGCAACTTGATGAT |
| CD45RA | HI100 | TCAATCCTTCCGCTT |
| CD194 (CCR4) | L291H4 | AGCTTACCTGCACGA |
| CD4 | RPA-T4 | TGTTCCCGCTCAACT |
| CD16 | 3G8 | AAGTTCACTCTTTGC |
| CD25 | BC96 | TTTGTCCTGTACGCC |
| CD45RO | UCHL1 | CTCCGAATCATGTTG |
| CD279 (PD-1) | EH12.2H7 | ACAGCGCCGTATTTA |
| TIGIT (VSTM3) | A15153G | TTGCTTACCGCCAGA |
| CD69 | FN50 | GTCTCTTGGCTTAAA |
| CD197 (CCR7) | G043H7 | AGTTCAGTCAACCGA |
| CD152 (CTLA-4) | BNI3 | ATGGTTCACGTAATC |
| CD27 | O323 | GCACTCCTGCATGTA |
| CD95 (Fas) | DX2 | CCAGCTCATTAGAGC |
| CD134 (OX40) | Ber-ACT35 (ACT35 | AACCCACCGTTGTTA |
| HLA-DR | L243 | AATAGCGAGCAAGTA |
| CD366 (Tim-3) | F38-2E2 | TGTCCTACCCAACTT |
| CD278 (ICOS) | C398.4A | CGCGCACCCATTAAA |
| CD39 | A1 | TTACCTGGTATCCGT |
| CD137 (4-1BB) | 4B4-1 | CAGTAAGTTCGGGAC |
| CD184 (CXCR4) | 12G5 | TCAGGTCCTTTCAAC |
| CD226 (DNAM-1) | 11A8 | TCTCAGTGTTTGTGG |
| CD28 | CD28.2 | TGAGAACGACCCTAA |
| CD45 | HI30 | TGCAATTACCCGGAT |
| CD103 (Integrin αE) | Ber-ACT8 | GACCTCATTGTGAAT |
| CD127 (IL-7Rα) | A019D5 | GTGTGTTGTCCTATG |

All antibodies from Biolegend.

**Table S7. TotalSeq-A antibodies used with 3'v3 scRNA-seq.**

| Specificity | Clone | Barcode Sequence |
|---|---|---|
| CD115 (CSF-1R) | 9-4D2-1E4 | AATCACGGTCCTTGT |
| CD117 (c-kit) | 104D2 | AGACTAATAGCTGAC |
| CD11b | ICRF44 | GACAAGTGATCTGCA |
| CD123 | 6H6 | CTTCACTCTGTCAGG |
| CD127 (IL-7Rα) | A019D5 | GTGTGTTGTCCTATG |
| CD13 | WM15 | TTTCAACGCCCTTTC |
| CD133 | S16016B | GTAAGACGCCTATGC |
| CD14 | 63D3 | CAATCAGACCTATGA |
| CD152 (CTLA-4) | BNI3 | ATGGTTCACGTAATC |
| CD16 | 3G8 | AAGTTCACTCTTTGC |
| CD184 (CXCR4) | 12G5 | TCAGGTCCTTTCAAC |
| CD19 | HIB19 | CTGGGCAATTACTCG |
| CD194 (CCR4) | L291H4 | AGCTTACCTGCACGA |
| CD197 (CCR7) | G043H7 | AGTTCAGTCAACCGA |
| CD2 | TS1/8 | TACGATTTGTCAGGG |
| CD22 | S-HCL-1 | GGGTTGTTGTCTTTG |
| CD223 (LAG-3) | 11C3C65 | CATTTGTCTGCCGGT |
| CD226 (DNAM-1) | 11A8 | TCTCAGTGTTTGTGG |
| CD244 (2B4) | C1.7 | TCGCTTGGATGGTAG |
| CD25 | BC96 | TTTGTCCTGTACGCC |
| CD26 | BA5b | GGTGGCTAGATAATG |
| CD274 (B7-H1, PD-L1) | 29E.2A3 | GTTGTCCGACAATAC |
| CD279 (PD-1) | EH12.2H7 | ACAGCGCCGTATTTA |
| CD3 | UCHT1 | CTCATTGTAACTCCT |
| CD32 | FUN-2 | GCTTCCGAATTACCG |
| CD33 | P67.6 | TAACTCAGGGCCTAT |
| CD34 | 581 | GCAGAAATCTCCCTT |
| CD366 (Tim-3) | F38-2E2 | TGTCCTACCCAACTT |
| CD38 | HIT2 | TGTACCCGCTTGTGA |
| CD4 | RPA-T4 | TGTTCCCGCTCAACT |
| CD44 | BJ18 | AATCCTTCCGAATGT |
| CD45 | HI30 | TGCAATTACCCGGAT |
| CD45RA | HI100 | TCAATCCTTCCGCTT |
| CD45RO | UCHL1 | CTCCGAATCATGTTG |
| CD56 (NCAM) | 5.1H11 | TCCTTTCCTGATAGG |
| CD69 | FN50 | GTCTCTTGGCTTAAA |
| CD7 | CD7-6B7 | TGGATTCCCGGACTT |
| CD8 | SK1 | GCGCAACTTGATGAT |
| CD90 (Thy1) | 5E10 | GCATTGTACGATTCA |
| CD95 (Fas) | DX2 | CCAGCTCATTAGAGC |
| CD96 (TACTILE) | NK92.39 | TGGCCTATAAATGGT |
| CD99 | 3B2/TA8 | ACCCGTCCCTAAGAA |
| CLEC12a | 50C1 | CATTAGAGTCTGCCA |
| HLA-A,B,C | W6/32 | TATGCGAGGCTTATC |
| HLA-DR | L243 | AATAGCGAGCAAGTA |
| HLA-DR,DP,DQ | Tu39 | AGCTACGAGCAGTAG |

All antibodies from Biolegend.

**Table S8. Oligos used in ADT cDNA and library amplifiation.**

| | |
|---|---|
| ADT additive primer | 5'CCTTGGCACCCGAGAATTCC |
| SI-PCR primer | 5'AATGATACGGCGACCACCGAGATCTACACTCTTTCCCTACACGACGCTC |
| Illumina small RNA primer RPI-2 | 5'CAAGCAGAAGACGGCATACGAGATACATCGGTGACTGGAGTTCCTTGGCACCCGAGAATTCCA |
| Illumina small RNA primer RPI-3 | 5'CAAGCAGAAGACGGCATACGAGATGCCTAAGTGACTGGAGTTCCTTGGCACCCGAGAATTCCA |
| Illumina small RNA primer RPI-5 | 5'CAAGCAGAAGACGGCATACGAGATCACTGTGTGACTGGAGTTCCTTGGCACCCGAGAATTCCA |
| Illumina small RNA primer RPI-7 | 5'CAAGCAGAAGACGGCATACGAGATGATCTGGTGACTGGAGTTCCTTGGCACCCGAGAATTCCA |
